# Predicting future cognitive impairment in preclinical Alzheimer’s disease using amyloid PET and MRI: a multisite machine learning study

**DOI:** 10.1101/2025.10.15.25337507

**Authors:** Braden Yang, Tom Earnest, Murat Bilgel, Marilyn S. Albert, Sterling C. Johnson, Christos Davatzikos, Guray Erus, Colin L. Masters, Susan M. Resnick, Michael I. Miller, Arnold Bakker, John C. Morris, Tammie L.S. Benzinger, Brian A. Gordon, Aristeidis Sotiras, the Alzheimer’s Disease Neuroimaging Initiative, the Preclinical Alzheimer’s Disease Consortium

## Abstract

Predicting the likelihood of developing Alzheimer’s disease (AD) dementia in at-risk individuals is important for the design of and optimal recruitment for clinical trials of disease-modifying therapies. Machine learning (ML) has been shown to excel in this task; however, there remains a lack of models developed specifically for the preclinical AD population, who display early signs of abnormal brain amyloidosis but remain cognitively unimpaired. Here, we trained and evaluated ML classifiers to predict whether individuals with preclinical AD will progress to mild cognitive impairment or dementia within multiple fixed time windows, ranging from one to five years. Models were trained on regional imaging features extracted from amyloid positron emission tomography and magnetic resonance imaging pooled across seven independent sites and from two amyloid radiotracers ([^18^F]-florbetapir and [^11^C]-Pittsburgh-compound-B). Out-of-sample generalizability was evaluated via a leave-one-site-out and leave-one-tracer-out cross-validation. Classifiers achieved an out-of-sample receiver operating characteristic area-under-the-curve of 0.66 or greater when applied to all except one hold-out sites and 0.72 or greater when applied to each hold-out radiotracer. Additionally, when applying our models in a retroactive cohort enrichment analysis on A4 clinical trial data, we observed increased statistical power of detecting differences in amyloid accumulation between placebo and treatment arms after enrichment by ML stratifications. As emerging investigations of new disease-modifying therapies for AD increasingly focus on asymptomatic, preclinical populations, our findings underscore the potential applicability of ML-based patient stratification for recruiting more homogeneous cohorts and improving statistical power for detecting treatment effects for future clinical trials.

**Highlights:**

- Machine learning can predict future cognitive impairment in preclinical Alzheimer’s
- Models achieved high out-of-sample ROC-AUC on external sites and PET tracers
- Models were able to distinguish cognitively stable from decliners in the A4 cohort
- ML cohort enrichment enhanced secondary treatment effect detection in the A4 cohort

## 1 Introduction

Alzheimer’s disease (AD) is a progressive neurodegenerative disease marked by the accumulation of amyloid-β plaques and tau neurofibrillary tangles in the brain, which leads to neuronal cell death and eventual development of cognitive deficits such as short-term memory loss, impairments in orientation, or difficulty with written and verbal communication. According to current models of the AD pathogenesis timeline, accumulation of these pathologies typically begin years before the onset of clinically-measurable symptoms (Jack and Holtzman, 2013; Jack Jr. et al., 2024). Thus, clinical trials of AD treatments have largely focused on disease-modifying therapies, which target these pathologies in the hopes of altering the biological course of the disease and slowing or even preventing the progression of clinical symptoms (Dyck et al., 2023; Sims et al., 2023). Yet to date, there have only been a handful of approved disease-modifying therapies, with the rest failing to demonstrate a significant treatment effect (Kim et al., 2022). A potential reason for this high failure rate is that recruited participants may have been at too advanced of a disease stage for treatments to meaningfully alter its progression (Sperling et al., 2014), with the majority of previously conducted trials primarily recruiting individuals who already displayed objective cognitive impairment (Huang et al., 2020; Sims et al., 2023; Swanson et al., 2021). In response, newly emerging trials have transitioned to targeting preclinical AD, where patients exhibit abnormal brain amyloidosis while showing no signs of cognitive impairment. Furthermore, failures may have also been attributed to high heterogeneity in the phenotypic expression of the disease, particularly in the rates of cognitive decline over time. This heterogeneity potentially leads to differential responses to administered therapies, which may negatively impact the estimation of treatment effects (Jutten et al., 2021).

To mitigate this issue, stratification models for distinguishing fast-cognitive decliners from slow or non-decliners are commonly employed in AD clinical trials. These models enable cohort enrichment by identifying and selectively enrolling candidates who are most likely to benefit from disease-modifying therapies in order to increase the likelihood of observing a potential treatment effect (Ballard et al., 2019; Wolz et al., 2016). Additionally, biased allocation of fast-decliners and slow-decliners into control and treatment groups may lead to overestimation or underestimation of a potential treatment effect. Proper stratification of clinical trial participants ensures that an equal distribution of fast-to-slow decliners are allocated into each trial arm and may aid in reducing uncertainties in treatment effect estimation, on top of reducing the required sample size and cost of running a trial (Wang et al., 2024). Previous trials have used single risk assessments such as neuropsychological testing and quantitative amyloid and tau measurements from positron emission tomography (PET) to stratify participants into high and low risk groups (Abdelnour et al., 2022; Sims et al., 2023; Swanson et al., 2021). However, recent evidence has suggested that data-driven stratification models built on machine learning (ML) can outperform single risk factor stratification in enhancing clinical trial operations (Birkenbihl et al., 2024; Gladstein et al., 2025; Nallapu et al., 2025; Tam et al., 2022; Wang et al., 2024). By leveraging information across multiple data modalities such as imaging, genetics and cognitive assessments, ML models can learn complex patterns within these multiple modalities to provide more accurate stratifications. These stratifications may be used to guide a more optimized cohort enrichment, leading to increased power of detect hypothetical treatment effects (Nallapu et al., 2025), reduction in random allocation bias (Wang et al., 2024), and reduction in the sample size required to observe a treatment effect (Birkenbihl et al., 2024; Gladstein et al., 2025; Tam et al., 2022). Among data modalities commonly used for stratification, medical imaging data like PET and magnetic resonance imaging (MRI) are particularly useful for probing the topographical distribution of core and associated AD pathologies in the brain (Jack Jr. et al., 2024), which have been shown to be highly discriminative of cognitive decliners from non-decliners (Mathotaarachchi et al., 2017; Pascoal et al., 2020; Pfeil et al., 2021).

Despite recent progress in developing ML models for this application, previous literature has predominantly focused on individuals who already exhibit cognitive deficits in the form of mild cognitive impairment (MCI) (Ezzati et al., 2021; Samper-González et al., 2018), with many studies only training on features extracted from structural MRI (Basaia et al., 2019; Mathotaarachchi et al., 2017; Misra et al., 2009). Efforts have been made to develop models for the cognitively unimpaired (CU) population that also incorporate amyloid PET, but most have either imposed a single time-to-progression window (Choi et al., 2024; Dansson et al., 2021; Pfeil et al., 2021) or don’t ensure the presence of amyloid pathology in their CU cohorts (Karaman et al., 2022). Additionally, with trials routinely involving large consortiums of institutions, each utilizing a different set of data acquisition protocols and scanner models, this introduces site-specific biases in the collected data which may negatively affect the ability of models to generalize on out-of-sample data. Furthermore, disparities in the pharmacokinetics and binding properties of different PET radiotracers lead to significant biases in quantitative amyloid PET measurements (Landau et al., 2014; Su et al., 2019; Villemagne et al., 2012). For these reasons, a rigorous evaluation of ML generalizability to external sites and different amyloid PET radiotracers is needed.

Motivated by these gaps, the current study seeks to develop and evaluate ML classifiers to predict future cognitive decline in asymptomatic, preclinical AD patients. We trained support vector machines (SVM) on regional imaging features extracted from amyloid PET and structural MRI to perform binary classification of whether an individual will develop cognitive deficits within multiple follow-up times, ranging from one year to five years. We obtained data from seven independent sites and two amyloid PET radiotracers and estimated the out-of-sample performance of trained models via leave-one-site-out and leave-one-tracer-out cross-validation. Lastly, to gauge the benefit of applying these models in a clinical trial setting, we utilized data from the Anti-Amyloid Treatment in Asymptomatic Alzheimer’s (A4) study (Sperling et al., 2023) to estimate the change in statistical power to detect treatment effects after retroactive cohort enrichment using our models’ predictions.

## 2 Materials and methods

### 2.1 Participants and data

Participants and their respective data were obtained from the following independent studies and consortia – the A4 study (Sperling et al., 2023), the Alzheimer’s Disease Neuroimaging Initiative (ADNI), the Harvard Aging Brain Study (HABS) (Dagley et al., 2017), the Mayo Clinic Study of Aging (MCSA) (Roberts et al., 2008), the Open Access Series of Imaging Studies 3 (OASIS) (LaMontagne et al., 2019), and the Preclinical Alzheimer’s Disease Consortium (PAC). ADNI, launched in 2003 as a public-private partnership and led by Principal Investigator Michael W. Weiner, MD, has aimed to test whether serial MRI, PET, other biological markers, and clinical and neuropsychological assessment can be combined to measure the progression of MCI and early AD. PAC is a consortium comprising five AD-focused datasets – the Adult Children Study (ACS) (Coats and Morris, 2005; Price and Morris, 1999), the Australian Imaging Biomarkers and Lifestyle (AIBL) study (Fowler et al., 2021), the Biomarkers of Cognitive Decline Among Normal Individuals (BIOCARD) study (Albert et al., 2014), the Baltimore Longitudinal Study of Aging (BLSA) (Resnick et al., 2000), and the Wisconsin Registry for Alzheimer’s Prevention (WRAP) (Johnson et al., 2018). All participants provided informed consent to participate in their respective studies in accordance with the Declaration of Helsinki, and ethics approvals were obtained by each site’s respective institutional review board. From the A4 study, only participants from the placebo arm of the clinical trial or from the Longitudinal Evaluation of Amyloid Risk and Neurodegeneration (LEARN) study were used to train the ML models. Participants from the Solanezumab treatment arm were excluded from training and were only used as an additional evaluation set (detailed in section *2.4 A4 trial analyses*). Additionally, we omitted ACS from our analyses since this dataset had significant participant overlap with OASIS.

All participants with available [^18^F]-florbetapir (FBP) or [^11^C]-Pittsburgh-compound-B (PiB) amyloid PET imaging were selected. We restricted these scans to be amyloid-positive, defined as having a mean cortical amyloid PET signal of greater than a threshold value. We used the dataset-specific definition of the summary cortical region mask and cutoff signal that was optimally determined for each dataset. Further details on how amyloid positivity was determined for each site can be found in the Supplementary Methods. All scans were matched with an accompanying structural T1-weighted MRI scan within one year of the PET acquisition date. Baseline age, sex and *APOE4* carriership were selected as additional predictors. We used the global Clinical Dementia Rating^®^ (CDR^®^) (Morris, 1997), which rates an individual’s cognitive performance in six domains (memory, orientation, judgment and problem solving, community affairs, home and hobbies, and personal care), as a measure of overall dementia severity. All PET scans were matched to a global CDR assessment within one year of the PET acquisition date. Participants who were missing any matching data were excluded from further analyses. The analysis was restricted to participants who were CU at the time of the amyloid PET scan, defined as a global CDR score of zero. We labeled participants as stable if they remained CU for at least five years from the time of the amyloid PET, and as progressor if they converted to a global CDR of greater than zero within a follow-up time that varied from one to five years. Progressors who reverted to CU were excluded. Additionally, stables who did not have a follow-up CDR of zero at least five years from the time of the amyloid PET were excluded. We observed a larger proportion of stable individuals compared to progressors; to reduce class imbalance, we imposed the five-year stability criterion on the stable cohort to obtain the most homogeneous group of participants, with presumably the least amount of pathology, to represent our control group. PAC did not provide CDR scores; we instead used the clinical diagnoses in place of CDR, with a diagnosis of either MCI or dementia being equivalent to CDR>0. In total, 343 stable and 247 progressor participants across seven sites (A4, ADNI, AIBL, BLSA, HABS, MCSA, OASIS) were selected for this study. We excluded BIOCARD and WRAP from our analyses due to too few participants meeting the selection criteria. Participant counts after exclusions are illustrated in Supplementary Fig. 1.

### 2.2 Image acquisition and processing

All scans underwent a standard PET-MRI processing pipeline consisting of segmentation of the MRI into regions-of-interest (ROI) and derivation of regional amyloid standardized uptake value ratios (SUVR) from the amyloid PET. Each participant’s T1-weighted MRI was preprocessed using a fully automated pipeline which included N4 bias correction (Tustison et al., 2010), skull stripping, and segmentation into 145 cortical and subcortical ROI using Multi-atlas region Segmentation utilizing Ensembles of registration algorithms and parameters (MUSE) (Doshi et al., 2016). Manual quality checks were performed on the outputs to ensure correctness of segmentations. We selected 125 relevant ROI and extracted their volumes, normalized by the total intracranial volume, to be used as the volumetric input features of our model.

FBP and PiB PET scans were downloaded either in a frame-averaged, smoothed and uniform resolution format (ADNI only) or as the full dynamic scan (all other datasets). For dynamic scans, we selected frames within the 50-70 minute window post-tracer injection for FBP and the 40-60 minute window for PiB. All frames were realigned to remove patient motion, averaged, and coregistered to the T1 image using FSL FLIRT rigid body registration (Smith et al., 2004). Additionally, we performed an iterative smoothing operation by applying a Gaussian smoothing kernel with progressively increasing full width at half maximum until a target resolution was reached (estimated using AFNI’s *3dFWHMx* tool). The target resolution was set to the *3dFWHMx*-estimated resolution of ADNI frame-averaged scans, which was found to be approximately 10mm isotropic. After smoothing, the PET image was converted to an SUVR image by dividing each voxel by the average signal of the left and right cerebellar gray matter regions. Finally, the MRI-to-PET transformation was applied to the anatomical MUSE ROI labels to sample them to native PET space, and regional SUVRs were computed by averaging the SUVR image over each ROI. For ADNI scans, we only performed coregistration to the T1, SUVR image computation and derivation of regional SUVRs. Manual quality checks were performed to ensure correctness of the frame alignment, PET-MRI coregistration and SUVR computation steps. Of the 145 total ROI, we selected 114 cortical and subcortical gray matter regions to be used as the amyloid input features of our model. The full list of ROIs used in this study are listed in Supplementary Table 1.

### 2.3 Model development and evaluation

#### 2.3.1 Training

The overview of the study is illustrated in Figure 1. We trained SVM binary classifiers with linear kernel to predict whether an individual will progress to CDR greater than zero using baseline amyloid PET SUVR, volumetric and non-imaging (i.e., age, sex, *APOE4*) features. We chose to train SVM classifiers since it has been widely used and extensively validated for AD diagnosis classification and prediction of future cognitive decline (Rathore et al., 2017). To account for imbalances in class frequency, we employed class-balanced weighting in which misclassifications of the minority class (progressor class in most cases) were more heavily penalized during optimization. Five separate time-to-progression models were trained independently of one another to predict progressor status, with the time window varied between one and five years. For example, the three-year model predicts if an individual will progress to a CDR of greater than zero within three years from baseline. Note that each selected time-to-progression would result in a different set of progressor participants. However, we used the same set of stable participants who remained CU for at least five years as the control group in all five models.

**Figure 1.**
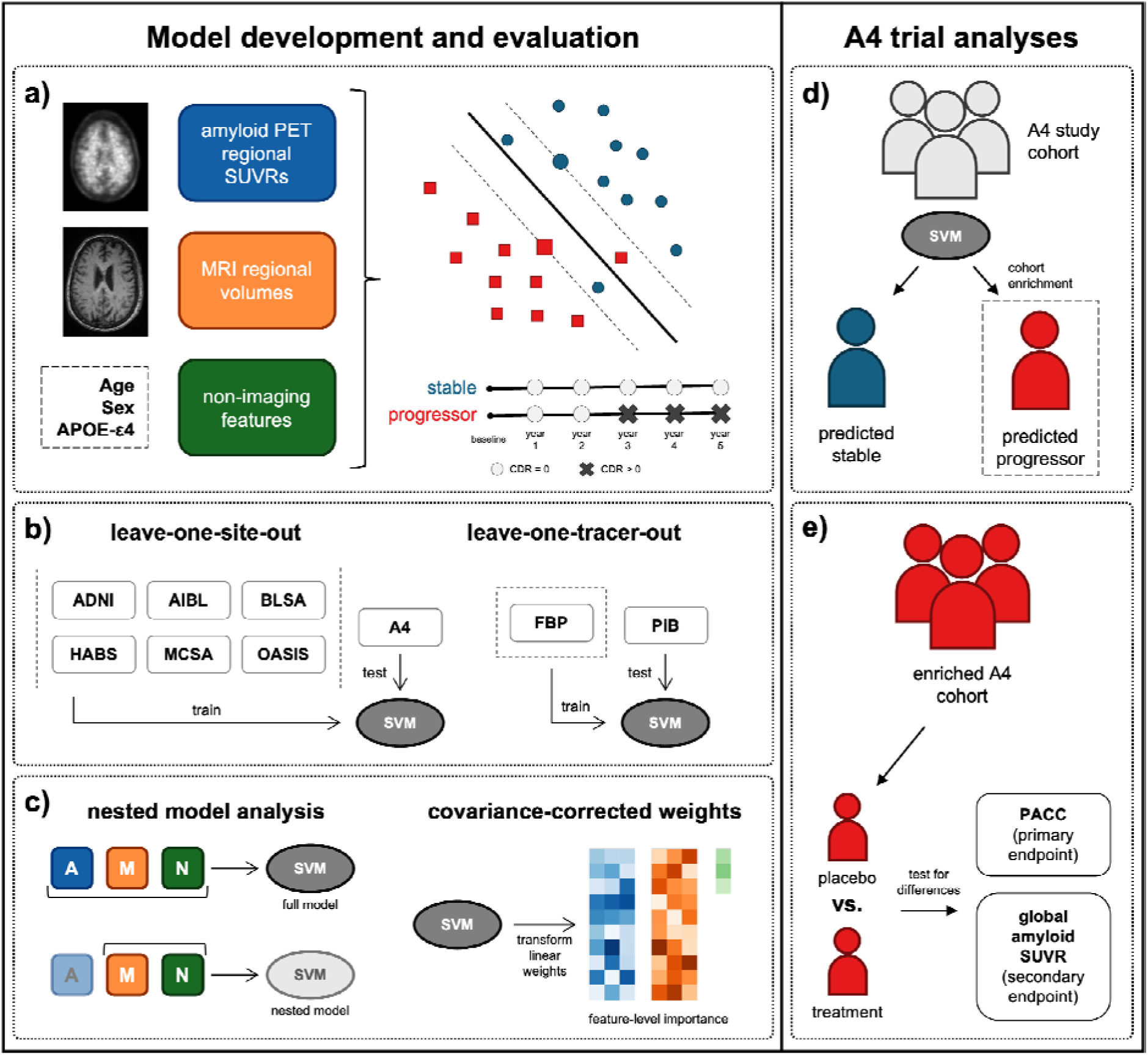
Overview of the study. Analyses were split into *model development and evaluation* (**a-c**) and *A4 trial analyses* (**d-e**). **(a)** Amyloid PET regional SUVRs, MRI regional volumes, and non-imaging features (age, sex, APOE-ε4) were inputted to train support vector machine classifiers to distinguish preclinical AD stables from progressors, where stables are individuals who remain CDR=0 for at least five years from baseline and progressors are individuals who convert to CDR>0 within a specified time window. **(b)** To evaluate model generalizability to unseen data, models were trained on all but one site or one tracer and tested on the hold-out site or tracer. **(c)** To estimate relative importance of feature modalities, nested models were trained on a subset of features and classification performance was compared to the model trained on the full feature set. To obtain individual feature-level importance, linear SVM weights were transformed using the data covariance matrix. **(d)** The SVM classifier trained on all sites except A4 was applied to the A4 cohort to predict progressor status. Only predicted progressors were retained to retroactively enrich the cohort. **(e)** To gauge the benefit of machine learning cohort enrichment, differences in PACC and global amyloid SUVR were tested between placebo and treatment arms post-enrichment. *Abbreviations*. CDR: Clinical Dementia Rating, PACC: Preclinical Alzheimer’s Cognitive Composite, SUVR: standardized uptake value ratio, SVM: support vector machine.

#### 2.3.2 Cross-validation

We employed a leave-one-site-out and leave-one-tracer-out cross-validation framework, in which data from one site or from one PET tracer were held out for independent testing, while the remaining data were used for training. Along with five times-to-progression, this resulted in 45 total models. All continuous features were z-scored using the standard deviation of only the CU-stable participants of the training fold, which was shown to enhance separation between linearly separable groups (Linn et al., 2016). A grid search with five-fold cross-validation was conducted using the training fold to determine the optimal value of hyperparameter C, which regulates how strongly misclassifications impact the SVM optimization. We optimized hyperparameter C independently for each of our 45 models. To evaluate model performance on the leave-out test set, the receiver operating characteristic area-under-the-curve (ROC-AUC), accuracy, balanced accuracy, F1 score, sensitivity, specificity, positive predictive value (PPV) and negative predictive value (NPV) were reported.

#### 2.3.3 Centiloid sensitivity analysis

To address biases in amyloid PET SUVRs due to the inclusion of multiple tracers, we performed a sensitivity analysis where we harmonized regional SUVRs into regional Centiloids before inputting as features to the SVM classifier. The Centiloid scale is a method of linearly transforming raw amyloid PET SUVRs into a scale that is standardized across different tracers (Klunk et al., 2015). We utilized the conversion equations that were uniquely calibrated to each site to convert SUVRs to Centiloid, then retrained our models with Centiloids as the amyloid PET features. Equations were only available for A4, ADNI, MCSA and OASIS, so we included only these sites in the sensitivity analysis. The DeLong test was used to compare ROC-AUC between models utilizing regional Centiloids versus regional SUVRs. Bonferroni correction was applied to p-values to adjust for multiple comparisons.

#### 2.3.4 Feature importance

We assessed the relative importance of each input feature for influencing the classification task via two methods. First, to estimate the relative importance of each modality of input feature (amyloid PET SUVR, MRI volume, or non-imaging features), we trained nested models wherein a reduced subset of modalities was used as input. We then computed the change in ROC-AUC on the hold-out test set relative to the full model as a measure of the relative contribution of that modality to the prediction. The DeLong test with Bonferroni correction for multiple comparisons was performed to test for statistical significance of ROC-AUC differences. Secondly, to derive a feature-level measure of the strength of association between each input feature and the progressor class, we computed the covariance-corrected linear SVM weights of the full model, as proposed by Haufe et al. (2014). This approach transforms classifier weights into interpretable estimates of the feature-level association with the progressor class, with high positive values indicating positive association, high negative values indicating negative association, and values close to zero indicating little to no association. A major advantage of this is that feature importance estimates are less prone to artificial distortion caused by collinearity of features, unlike impurity-based metrics for models such as random forest (Strobl et al., 2008). For each classifier, we applied this method to estimate importance values of each input feature, and we averaged the values across all 45 classifiers to derive an overall feature importance. Additionally, to gauge how the magnitude of feature importance varies in relation to the time-to-progression, we estimated the slope of the absolute value of feature importance versus time-to-progression via an ordinary least squares linear regression.

### 2.4 A4 trial analyses

To further validate trained SVM classifiers, we performed three analyses using the extended A4 study cohort as the external testing set. This extended cohort consisted of participants in the placebo arm and participants treated with Solanezumab, an immunoglobulin monoclonal antibody designed to increase amyloid clearance in the brain. Firstly, we applied the classifier trained on all datasets except A4 to classify A4 participants into stables and progressors using imaging acquired during the screening phase of the trial. We then compared the Preclinical Alzheimer’s Cognitive Composite (PACC), a composite score which has been shown to better track early AD-related cognitive changes (Donohue et al., 2014), between predicted stables and progressors. Mirroring previous work (Donohue et al., 2023; Sperling et al., 2023), we fitted a natural cubic spline model on longitudinal PACC scores as the target variable, using continuous time-from-baseline as the independent variable and baseline age, years of education, *APOE4* carriership, PACC test version administered, and baseline FBP SUVR of a cortical composite region as covariates. We then estimated a model-adjusted mean PACC score and 95% confidence interval at the 240-week follow-up point (corresponding to the end of the placebo controlled study period) and compared the estimates derived for predicted stables versus predicted progressors using a two-sample t-test. This analysis was independently performed within the placebo and Solanezumab arms.

Secondly, we applied the SVM classifications to conduct retroactive enrichment wherein we restricted the cohort to only include predicted progressors. We then tested for treatment effects post-enrichment in the primary endpoint (model-adjusted PACC) and a secondary endpoint (change in amyloid PET SUVR). For the primary endpoint, we fitted the same natural cubic spline model with covariates on the longitudinal PACC scores of the enriched placebo and treatment cohorts. Model-adjusted means and 95% confidence intervals of PACC at the 240-week follow-up point were estimated and compared between placebo and Solanezumab arms using a two-sample t-test. For the secondary endpoint, we computed the change in FBP cortical SUVR from baseline to the 240-week follow-up point and used this metric as the target variable. An analysis of covariance (ANCOVA) was fitted to compare the mean change in SUVR between placebo and Solanezumab arms, controlling for baseline age, *APOE4* carriership and baseline SUVR as covariates.

Lastly, we performed a bootstrap analysis to estimate the statistical power of detecting a difference in the change in FBP cortical SUVR between baseline and 240-weeks, testing different sample sizes and sampling populations (i.e. enriched vs. unenriched). Here, we focused only on the secondary endpoint rather than the PACC primary endpoint since no statistically significant treatment effect was reported for the primary endpoint in the original A4 study. For a fixed sample size *n*, we randomly selected *n* participants each from the placebo and Solanezumab arms. The pool of participants to select from was either the full, unenriched cohort or the ML-enriched cohort. We then reran the ANCOVA as conducted previously to test for significant differences in the adjusted change in SUVR between the randomly selected cohorts. We repeated this procedure 1000 times. Power was estimated as the percentage of significant findings across all 1000 iterations. Samples sizes per trial cohort were varied from 10 to 80 in increments of 10.

## 3 Results

### 3.1 Cohort characteristics

Table 1 summarizes the cohort characteristics of each site and of the pooled data; p-values were derived from two-sample t-tests for all continuous variables and Fisher’s exact test for all categorical variables. Across all sites, progressors were older on average (75.9 ± 6.4) than stables (72.4 ± 5.6, *p < 0.001*) and displayed similar proportions of females-to-males and *APOE4* carriers-to-noncarriers compared to stables (*p > 0.05*). Progressors had a mean time to progression of 2.4 ± 1.3 years and stables remained CU for an average of 7.1 ± 1.6 years. Stables were scanned with a roughly equal proportion of FBP to PiB radiotracer, whereas progressors were more often scanned with FBP (60%). Within individual sites, only age in the A4 cohort displayed a statistically significant difference between stables and progressors across all demographics reported (*p < 0.001*).

**Table 1.** Cohort characteristics of stables and progressors from all individual sites and the pooled dataset.

|  | A4 |  | ADNI |  | AIBL |  | BLSA |  | HABS |  | MCSA |  | OASIS |  | pooled |  |
| --- | --- | --- | --- | --- | --- | --- | --- | --- | --- | --- | --- | --- | --- | --- | --- | --- |
|  | stable | progr<br>essor | stable | progr<br>essor | stable | progr<br>essor | stable | progr<br>essor | stable | progr<br>essor | stable | progr<br>essor | stable | progr<br>essor | stable | progr<br>essor |
| number of participants | 128 | 111 | 22 | 29 | 28 | 14 | 13 | 11 | 4 | 15 | 94 | 43 | 54 | 24 | 343 | 247 |
| age, mean (SD) | 70.05 (3.47) | 73.47 (5.17) | 74.25 (6.79) | 78.14 (6.00) | 72.14 (5.78) | 74.23 (6.83) | 77.08 (7.04) | 83.01 (4.77) | 75.56 (1.66) | 76.28 (5.33) | 75.43 (5.34) | 79.19 (7.09) | 70.75 (6.17) | 75.91 (6.68) | 72.41 (5.62) | 75.89 (6.43) |
| female, count (%) | 91 (71) | 63 (57) | 13 (59) | 18 (62) | 13 (46) | 5 (36) | 6 (46) | 3 (27) | 1 (25) | 13 (87) | 45 (48) | 20 (47) | 38 (70) | 11 (46) | 207 (60) | 133 (54) |
| APOE4 carriership, count (%) | 78 (61) | 63 (57) | 6 (27) | 11 (38) | 12 (43) | 10 (71) | 2 (15) | 5 (45) | 2 (50) | 9 (60) | 38 (40) | 26 (60) | 29 (54) | 12 (50) | 167 (49) | 136 (55) |
| amyloid PET tracer, count (%) |  |  |  |  |  |  |  |  |  |  |  |  |  |  |  |  |
| FBP | 128 (100) | 111 (100) | 22 (100) | 29 (100) | - | - | - | - | - | - | - | - | 20 (37) | 10 (42) | 170 (50) | 150 (61) |
| PIB | - | - | - | - | 28 (100) | 14 (100) | 13 (100) | 11 (100) | 4 (100) | 15 (100) | 94 (100) | 43 (100) | 34 (63) | 14 (58) | 173 (50) | 97 (39) |
| years to CDR>0 progression, mean (SD) | - | 2.35 (1.23) | - | 2.37 (1.26) | - | 3.04 (1.37) | - | 2.99 (1.34) | - | 2.75 (1.54) | - | 2.27 (1.05) | - | 2.42 (1.49) | - | 2.44 (1.27) |
| years of CU-stable, mean (SD) | 6.68 (0.90) | - | 7.55 (1.74) | - | 8.68 (2.50) | - | 7.31 (2.21) | - | 5.38 (0.32) | - | 6.84 (1.31) | - | 7.61 (1.97) | - | 7.10 (1.61) | - |
| years to CDR>0 progression, count (%) |  |  |  |  |  |  |  |  |  |  |  |  |  |  |  |  |
| 0-1 year | - | 5 (4.5) | - | 6 (21) | - | - | - | - | - | 3 (20) | - | 4 (9.3) | - | 7 (29) | - | 25 (10) |
| 1-2 years | - | 37 (33) | - | 6 (21) | - | 4 (29) | - | 4 (36) | - | 4 (27) | - | 13 (30) | - | 2 (8.3) | - | 70 (28) |
| 2-3 years | - | 33 (30) | - | 8 (28) | - | 2 (14) | - | 1 (9.1) | - | 1 (6.7) | - | 17 (40) | - | 8 (33) | - | 70 (28) |
| 3-4 years | - | 20 (18) | - | 4 (14) | - | 3 (21) | - | 2 (18) | - | 4 (27) | - | 8 (19) | - | 2 (8.3) | - | 43 (17) |
| 4-5 years | - | 16 (14) | - | 5 (17) | - | 5 (36) | - | 4 (36) | - | 3 (20) | - | 1 (2.3) | - | 5 (21) | - | 39 (16) |

\* $p < 0.05$
\*\* $p < 0.01$
\*\*\* $p < 0.001$

### 3.2 Model evaluation

We evaluated the external site generalizability of the SVM classifier using leave-one-site-out cross-validation. The classifier generalized well to most leave-out sites and times-to-progression, achieving an ROC-AUC of 0.66 or greater, with a majority exceeding 0.75 or above (Figure 2, Table 2). The exception was HABS, for which the ROC-AUC fell below 0.50. We also computed aggregate binary classification metrics by combining the out-of-sample predictions of every model on their respective leave-out site and comparing them to the combined ground truth labels from all sites (Table 2). Classifiers achieved an aggregate accuracy of >0.69 and balanced accuracy of >0.58. Classifiers were also highly specific (>0.74) and had high negative predictive value (>0.74). While models also achieved relatively high sensitivity, PPV and F1 score, these metrics were particularly sensitive to time-to-progression. Notably, smaller times-to-progression resulted in poorer sensitivity, PPV and F1 score, likely due to the fact that classifiers were trained on a fewer number of positive cases and were prone to overfitting to the control cases. Classifiers trained on higher times-to-progression exhibited a better balance between sensitivity/PPV to specificity/NPV.

**Figure 2.**
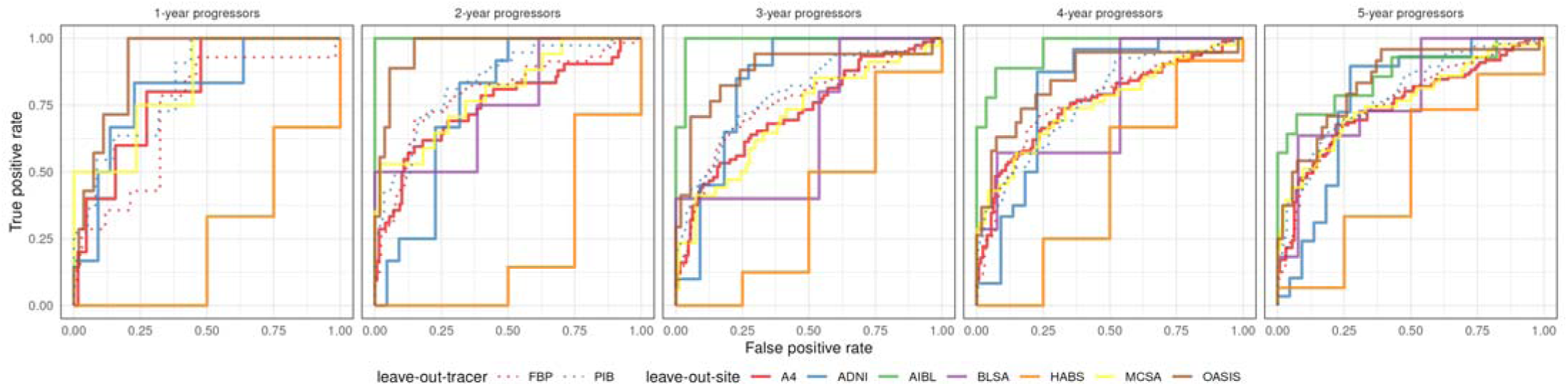
Receiver operating characteristic curves of SVM classifiers trained on each of five times-to-progression. Curves were obtained by applying each trained model to data from its respective hold-out group. Solid lines represent leave-out sites and dotted lines represent leave-out tracers. Across all times-to-progression, SVM classifiers achieved area-under-the-curve of 0.66 or greater in all leave-out sites except for HABS and 0.72 or greater in both leave-out tracers.

**Table 2.** Evaluation metrics of SVM classifiers of all five times-to-progression, tested on each site and tracer as the independent testing set.

|  | progression<br>time-horizon<br>(yrs) | leave-out site/tracer |  |  |  |  |  |  |  |  |  |  |
| --- | --- | --- | --- | --- | --- | --- | --- | --- | --- | --- | --- | --- |
|  |  | A4 | ADNI | AIBL | BLSA | HABS | MCSA | OASIS | aggregated<br>(site) | FBP | PIB | aggregated<br>(tracer) |
| Receiver<br>Operating<br>Characteris<br>tic Area-under-the-curve<br>(ROC-AUC) | 1 | 0.8062 | 0.8030 | - | - | 0.2500 | 0.8298 | 0.9074 | - | 0.7235 | 0.8282 | - |
|  | 2 | 0.7487 | 0.7576 | 1.000 | 0.7500 | 0.2143 | 0.8004 | 0.9630 | - | 0.7897 | 0.8352 | - |
|  | 3 | 0.7230 | 0.8364 | 0.9881 | 0.6615 | 0.3750 | 0.7078 | 0.8758 | - | 0.7537 | 0.7918 | - |
|  | 4 | 0.7573 | 0.8049 | 0.9603 | 0.7473 | 0.4583 | 0.7538 | 0.8489 | - | 0.7647 | 0.7679 | - |
|  | 5 | 0.7478 | 0.7743 | 0.8622 | 0.7902 | 0.5000 | 0.7640 | 0.8395 | - | 0.7569 | 0.7904 | - |
| Accuracy | 1 | 0.8872 | 0.8214 | 0.9286 | 0.5385 | 0.2857 | 0.6837 | 0.9016 | 0.8098 | 0.9239 | 0.6630 | 0.7935 |
|  | 2 | 0.7941 | 0.7353 | 0.8125 | 0.5294 | 0.2727 | 0.6126 | 0.8571 | 0.7306 | 0.7807 | 0.6095 | 0.6986 |
|  | 3 | 0.6897 | 0.8095 | 0.8235 | 0.5000 | 0.5000 | 0.6719 | 0.8310 | 0.7126 | 0.6960 | 0.6553 | 0.6772 |
|  | 4 | 0.7265 | 0.7826 | 0.8108 | 0.6000 | 0.5000 | 0.6471 | 0.8082 | 0.7169 | 0.6835 | 0.6102 | 0.6497 |
|  | 5 | 0.7155 | 0.7451 | 0.7619 | 0.6667 | 0.5263 | 0.6204 | 0.7692 | 0.6983 | 0.6469 | 0.6407 | 0.6441 |
| Balanced<br>Accuracy | 1 | 0.6531 | 0.5833 | 0.9286 | 0.5385 | 0.2500 | 0.7154 | 0.6336 | 0.5827 | 0.5328 | 0.6931 | 0.5925 |
|  | 2 | 0.6953 | 0.7197 | 0.8929 | 0.6923 | 0.3214 | 0.6990 | 0.8704 | 0.6948 | 0.6144 | 0.7205 | 0.6325 |
|  | 3 | 0.6601 | 0.8114 | 0.8929 | 0.5308 | 0.5000 | 0.6264 | 0.7881 | 0.6865 | 0.6162 | 0.7038 | 0.6273 |
|  | 4 | 0.7061 | 0.7822 | 0.8373 | 0.6593 | 0.5000 | 0.6722 | 0.7510 | 0.7045 | 0.6419 | 0.6810 | 0.6316 |
|  | 5 | 0.7051 | 0.7429 | 0.7500 | 0.6853 | 0.5167 | 0.6540 | 0.7176 | 0.6901 | 0.6280 | 0.6812 | 0.6270 |
| F1 Score | 1 | 0.2105 | 0.2857 | - | - | 0 | 0.1622 | 0.4000 | 0.1860 | 0.1250 | 0.2051 | 0.1915 |
|  | 2 | 0.5455 | 0.6400 | 0.5714 | 0.5000 | 0.2000 | 0.3944 | 0.6400 | 0.5042 | 0.3902 | 0.4459 | 0.4261 |
|  | 3 | 0.5655 | 0.8095 | 0.6667 | 0.4000 | 0.5714 | 0.4615 | 0.6667 | 0.5805 | 0.4196 | 0.5525 | 0.4938 |
|  | 4 | 0.6391 | 0.7917 | 0.6957 | 0.6000 | 0.6000 | 0.5636 | 0.6316 | 0.6355 | 0.4891 | 0.5892 | 0.5459 |
|  | 5 | 0.6458 | 0.7719 | 0.6667 | 0.7143 | 0.6400 | 0.5517 | 0.6087 | 0.6397 | 0.4645 | 0.6226 | 0.5513 |
| Sensitivity | 1 | 0.4000 | 0.1667 | - | - | 0 | 0.7500 | 0.2857 | 0.3200 | 0.0714 | 0.7273 | 0.3600 |
|  | 2 | 0.5000 | 0.6667 | 1.000 | 1.000 | 0.1429 | 0.8235 | 0.8889 | 0.6316 | 0.2759 | 0.8919 | 0.5158 |
|  | 3 | 0.5467 | 0.8500 | 1.000 | 0.6000 | 0.5000 | 0.5294 | 0.7059 | 0.6121 | 0.2913 | 0.8065 | 0.4848 |
|  | 4 | 0.5684 | 0.7917 | 0.8889 | 0.8571 | 0.5000 | 0.7381 | 0.6316 | 0.6538 | 0.3543 | 0.8765 | 0.5577 |
|  | 5 | 0.5586 | 0.7586 | 0.7143 | 0.9091 | 0.5333 | 0.7442 | 0.5833 | 0.6397 | 0.3267 | 0.8247 | 0.5223 |
| Specificity | 1 | 0.9062 | 1.000 | 0.9286 | 0.5385 | 0.5000 | 0.6809 | 0.9815 | 0.8455 | 0.9941 | 0.6590 | 0.8251 |
|  | 2 | 0.8906 | 0.7727 | 0.7857 | 0.3846 | 0.5000 | 0.5745 | 0.8519 | 0.7580 | 0.9529 | 0.5491 | 0.7493 |
|  | 3 | 0.7734 | 0.7727 | 0.7857 | 0.4615 | 0.5000 | 0.7234 | 0.8704 | 0.7609 | 0.9412 | 0.6012 | 0.7697 |
|  | 4 | 0.8438 | 0.7727 | 0.7857 | 0.4615 | 0.5000 | 0.6064 | 0.8704 | 0.7551 | 0.9294 | 0.4855 | 0.7055 |
|  | 5 | 0.8516 | 0.7273 | 0.7857 | 0.4615 | 0.5000 | 0.5638 | 0.8519 | 0.7405 | 0.9294 | 0.5376 | 0.7318 |
| Positive<br>Predictive<br>Value<br>(PPV) | 1 | 0.1429 | 1.000 | - | - | 0 | 0.0909 | 0.6667 | 0.1311 | 0.5000 | 0.1194 | 0.1304 |
|  | 2 | 0.6000 | 0.6154 | 0.4000 | 0.3333 | 0.3333 | 0.2593 | 0.5000 | 0.4196 | 0.6667 | 0.2973 | 0.3630 |
|  | 3 | 0.5857 | 0.7727 | 0.5000 | 0.3000 | 0.6667 | 0.4091 | 0.6316 | 0.5519 | 0.7500 | 0.4202 | 0.5031 |
|  | 4 | 0.7297 | 0.7917 | 0.5714 | 0.4615 | 0.7500 | 0.4559 | 0.6316 | 0.6182 | 0.7895 | 0.4438 | 0.5346 |
|  | 5 | 0.7654 | 0.7857 | 0.6250 | 0.5882 | 0.8000 | 0.4384 | 0.6364 | 0.6397 | 0.8033 | 0.5000 | 0.5837 |
| Negative<br>Predictive<br>Value<br>(NPV) | 1 | 0.9748 | 0.8148 | 1.000 | 1.000 | 0.4000 | 0.9846 | 0.9138 | 0.9446 | 0.9286 | 0.9744 | 0.9465 |
|  | 2 | 0.8444 | 0.8095 | 1.000 | 1.000 | 0.2500 | 0.9474 | 0.9787 | 0.8814 | 0.7941 | 0.9596 | 0.8482 |
|  | 3 | 0.7444 | 0.8500 | 1.000 | 0.7500 | 0.3333 | 0.8095 | 0.9038 | 0.8031 | 0.6867 | 0.8966 | 0.7564 |
|  | 4 | 0.7248 | 0.7727 | 0.9565 | 0.8571 | 0.2500 | 0.8382 | 0.8704 | 0.7825 | 0.6583 | 0.8936 | 0.7246 |
|  | 5 | 0.6899 | 0.6957 | 0.8462 | 0.8571 | 0.2222 | 0.8281 | 0.8214 | 0.7405 | 0.6100 | 0.8455 | 0.6802 |

We additionally performed leave-one-tracer-out validation to assess the generalizability of the SVM classifier to unseen tracers. The classifier achieved ROC-AUC of >0.72 across all times-to-progression for both leave-out tracers (Table 2). However, the model trained on PiB and tested on FBP had a low sensitivity and high specificity while the opposite was true for the model trained on FBP and tested on PiB. This may reflect bias in regional SUVRs between FBP and PiB tracers. To address this, we retrained the classifiers using Centiloid-harmonized regional amyloid PET SUVRs and reported the resulting changes in performance (Supplementary Table 2). After harmonization, models achieved a better balance of sensitivity to specificity for both leave-out tracers: sensitivity increased when testing on FBP, and specificity increased when testing on PiB. This was also accompanied by increased aggregate balanced accuracy across all times-to-progression. ROC-AUC was slightly decreased in some cases, but none of these changes were statistically significant via the DeLong test (*p > 0.05*).

### 3.3 Feature importance

The performances of nested models on each hold-out site are shown in Figure 3 and Supplementary Table 3. When amyloid PET was omitted from the input set, decreases of up to 0.223 in ROC-AUC were observed particularly at time windows of three years or greater (with the exception of A4), although these differences were not statistically significant (*p > 0.05*).

**Figure 3.**
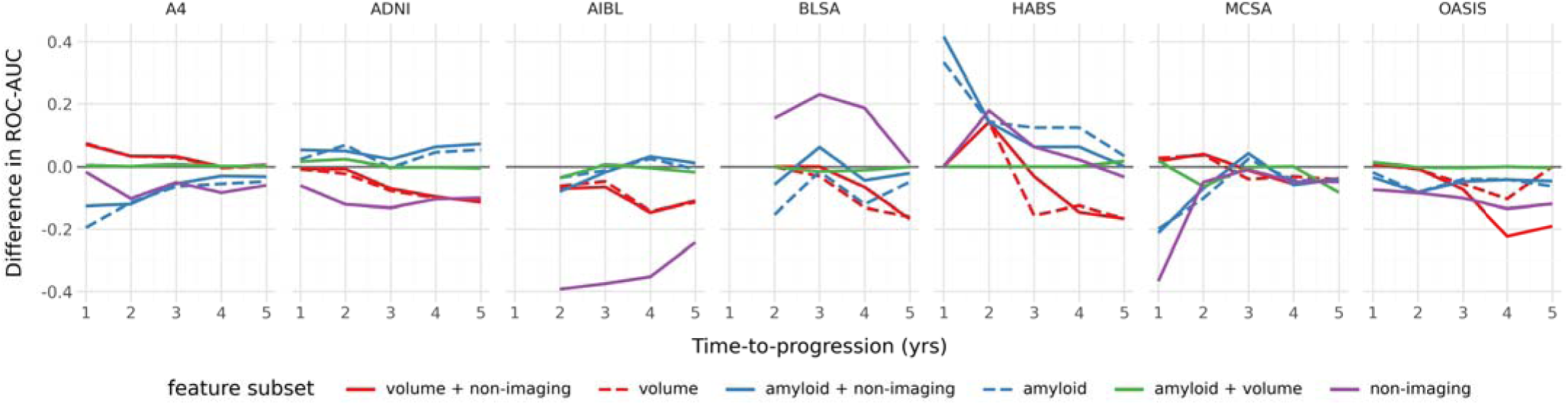
Difference in hold-out ROC-AUC of nested models relative to the full model trained on all feature modalities. Dotted lines represent imaging models that omit non-imaging features. In many hold-old sites, omission of amyloid PET features (red) at high times-to-progression led to a decrease in ROC-AUC compared to the full model. Omission of volumetric features (blue) resulted in less consistent trends. Except for BLSA and HABS, the non-imaging only model (purple) performs worse than the full model for nearly all times-to-progression. Omission of non-imaging features (green) led to nearly identical performance compared to the full model.

When MRI volumetric features were omitted from the input set, we observed less consistent trends across leave-out sites. For some sites such as A4 and OASIS, slight decreases in ROC-AUC were observed, whereas for other sites such as ADNI and HABS slight increases were observed. All differences were not statistically significant (*p > 0.05*). In many cases, the reduction of model generalizability when amyloid features were omitted was greater than when volumetric features were omitted, especially at later time windows, suggesting a higher relative importance of amyloid PET in these tasks. In all leave-out sites except for BLSA and HABS, the non-imaging only model consistently performed worse than the model integrating features from both imaging modalities, with the difference in ROC-AUC of the one-year model on MCSA being statistically significant (*p < 0.05*). Finally, omission of non-imaging features led to nearly identical performance compared to the full model.

The average importance of each input feature across all trained models is visualized in Figure 4 and Supplementary Fig. 2. For amyloid SUVR features, widespread contributions for predicting the progressor class were observed throughout the cortex, with bilateral inferior and middle temporal gyri exhibiting the strongest mean importance, followed by middle frontal, superior frontal and middle occipital gyri. In contrast, subcortical amyloid SUVR generally had much lower importance. For volumetric features, bilateral inferior lateral ventricular volumes had the highest absolute value of feature importance out of all features (including amyloid SUVR and non-imaging variables). This was followed by lateral ventricle, amygdala and hippocampus volume. Ventricular volumes all had positive importance values, whereas all other volumetric features had negative importances. Among the three non-imaging features, baseline age was most strongly associated with the progressor class. Neither sex nor *APOE4* carriership showed strong associations.

**Figure 4.**
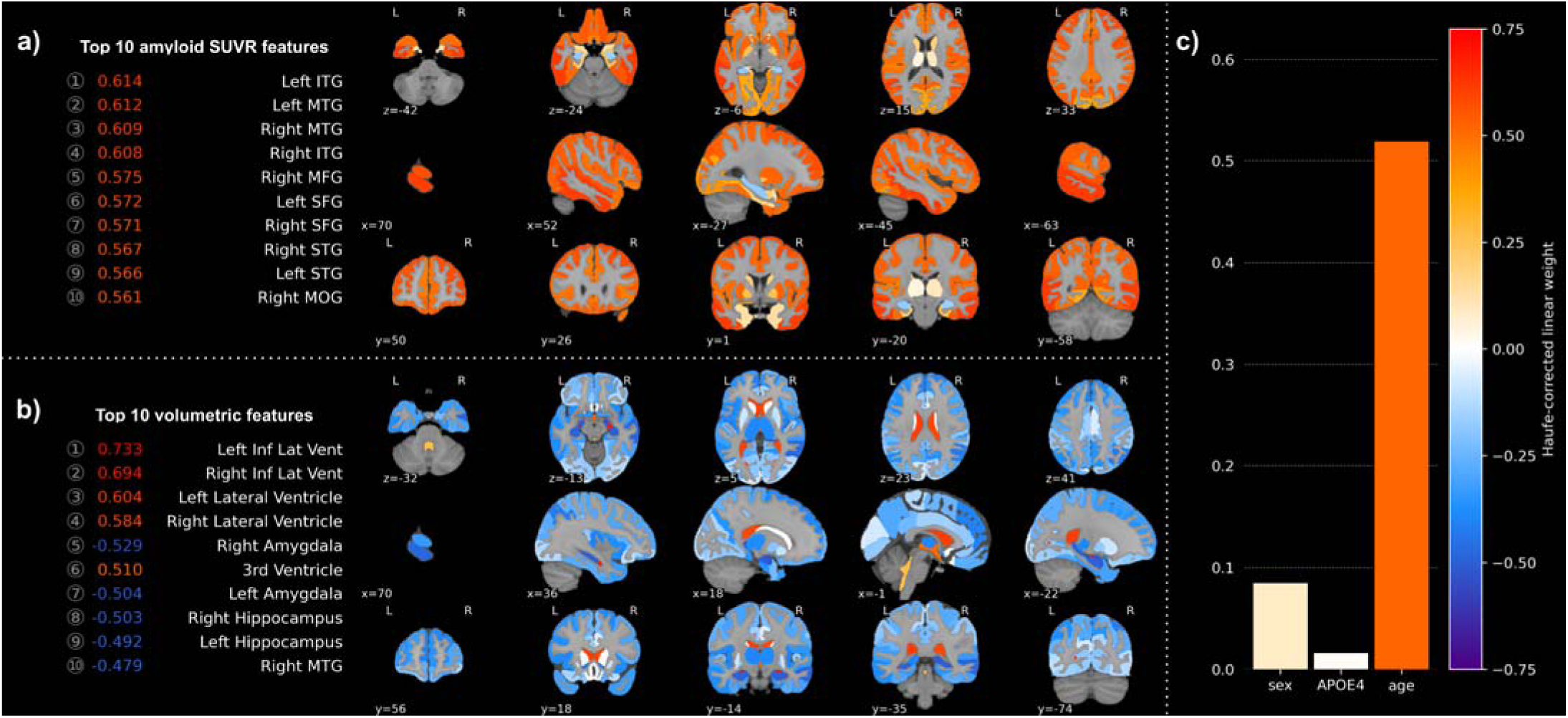
Covariance-corrected linear SVM weights per amyloid SUVR (**a**), volumetric (**b**), and non-imaging (**c**) feature. The values shown are the mean importance across all 45 trained models. The ten features with the highest magnitude of mean importance within each imaging modality are shown on the left. Among amyloid SUVR features, temporal, middle & superior frontal and middle occipital gyri had the highest magnitude mean importance. Among volumetric features, lateral ventricle, hippocampus and amygdala had the highest magnitude mean importance. Age was the only non-imaging feature with a high mean importance. Abbreviations. Inf Lat Vent: inferior lateral ventricle, ITG: inferior temporal gyrus, MFG: middle frontal gyrus, MOG: middle occipital gyrus, MTG: middle temporal gyrus, SFG: superior frontal gyrus, STG: superior temporal gyrus

We also estimated the rate-of-change of the absolute value of feature importance versus the time-to-progression via a linear regression (Supplementary Fig. 3 & 4). For amyloid SUVR features, regions in the orbitofrontal, temporal and occipital cortices had the highest rate of increase in importance as the time-to-progression window grew. Among volumetric regions, the right lateral inferior ventricles, right frontal operculum and bilateral posterior cingulate gyri had the highest rate of increase. Age had a modest rate of increase, while sex and *APOE4* did not show substantial change. Amyloid SUVR features displayed much greater rates-of-change in feature importance compared to either volumetric features or non-imaging features, suggesting that their influence on model outputs becomes more pronounced as the time-to-progression window expands to capture earlier disease progression. This was also consistent with the trends observed in the nested model analysis.

### 3.4 A4 trial analysis

We conducted retroactive analyses on the A4 clinical trial dataset to further validate our SVM classifiers and demonstrate their potential benefit for clinical trial cohort enrichment. Table 3 summarizes the characteristics of the unenriched A4 cohort and the retroactively enriched cohorts using each of our five time-to-progression models. In both placebo and Solanezumab arms, significant differences between the unenriched and enriched cohorts were observed in baseline age, baseline cortical FBP SUVR (except for the one-year progressors of the Solanezumab arm) and baseline PACC (except for the three-year progressors of the placebo arm) (*p < 0.05*). A lower percentage of females were selected for the enriched cohort in both trial arms compared to the unenriched cohort; this difference was only significant for the one-year progressors (*p < 0.01*). No significant differences were observed in *APOE4* carriership or annualized PACC rate-of-change (*p > 0.05*).

**Table 3.** Cohort characteristics of the placebo and Solanezumab treatment arms of the A4 study. The original, unenriched cohort and ML-enriched cohorts are shown.

|  | cohort |  |  |  |  |  |
| --- | --- | --- | --- | --- | --- | --- |
|  | original cohort | 1-year progressor | 2-year progressor | 3-year progressor | 4-year progressor | 5-year progressor |
| **Placebo** |  |  |  |  |  |  |
| number of participants | 444 | 72 | 132 | 158 | 149 | 150 |
| participants with FBP PET at 240-week follow-up, count (%) | 325 (73) | 43 (60) | 79 (60) | 108 (68) | 94 (63) | 95 (63) |
| baseline age, mean (SD) | 72.0 (5.0) | 75.2 (5.5)*** | 75.3 (5.2)*** | 74.6 (5.4)*** | 75.2 (5.2)*** | 74.9 (5.3)*** |
| female, count (%) | 270 (61) | 25 (35)** | 60 (45) | 86 (54) | 74 (50) | 78 (52) |
| APOE4 carriership, count (%) | 253 (57) | 46 (64) | 78 (59) | 90 (57) | 85 (57) | 87 (58) |
| baseline cortical FBP SUVR, mean (SD) | 1.36 (0.17) | 1.47 (0.20)** | 1.49 (0.18)*** | 1.46 (0.18)*** | 1.48 (0.18)*** | 1.49 (0.18)*** |
| baseline PACC, mean (SD) | 0.0 (2.6) | -1.5 (3.1)* | -1.5 (2.9)*** | -1.0 (3.1) | -1.3 (2.9)*** | -1.2 (3.0)** |
| annualized PACC rate-of-change, mean (SD) | -0.25 (1.24) | -0.48 (1.44) | -0.49 (1.51) | -0.49 (1.28) | -0.48 (1.45) | -0.48 (1.40) |
| **Solanezumab** |  |  |  |  |  |  |
| number of participants | 404 | 61 | 110 | 140 | 150 | 143 |
| participants with FBP PET at 240-week follow-up, count (%) | 305 (75) | 41 (67) | 74 (67) | 103 (74) | 107 (71) | 100 (70) |
| baseline age, mean (SD) | 72.2 (4.5) | 74.6 (5.1)* | 75.1 (4.4)*** | 74.3 (4.6)*** | 74.5 (4.5)*** | 74.6 (4.5)*** |
| female, count (%) | 227 (56) | 18 (30)** | 56 (51) | 74 (53) | 75 (50) | 71 (50) |
| APOE4 carriership, count (%) | 254 (63) | 39 (64) | 71 (65) | 93 (66) | 98 (65) | 90 (63) |
| baseline cortical FBP SUVR, mean (SD) | 1.37 (0.16) | 1.44 (0.17) | 1.51 (0.17)*** | 1.46 (0.18)*** | 1.48 (0.17)*** | 1.49 (0.17)*** |
| baseline PACC, mean (SD) | 0.05 (2.77) | -1.53 (2.89)** | -1.25 (2.57)*** | -0.96 (2.83)* | -1.01 (2.79)** | -1.11 (2.74)** |
| annualized PACC rate-of-change, mean (SD) | -0.31 (1.17) | -0.76 (1.43) | -0.78 (1.48) | -0.71 (1.37) | -0.69 (1.40) | -0.73 (1.45) |

\* $p < 0.05$
\*\* $p < 0.01$
\*\*\* $p < 0.001$

To validate that the SVM classifier accurately distinguish cognitive stables from decliners, we fitted a natural cubic spline model to longitudinal PACC scores of predicted stables and progressors and estimated the model-adjusted mean PACC score at the 240-week follow-up. In all cases except for the one-year progressor model in the Solanezumab arm, predicted stables had a significantly higher mean estimate of the model-adjusted PACC score at 240 weeks than predicted progressors (*p < 0.01*) (Figure 5, Supplementary Table 4). When using the five-year model, which roughly aligns with the duration of the placebo-controlled period of the A4 trial, predicted progressors of the Solanezumab arm had a mean adjusted PACC score of -3.92 ± 0.57, which was significantly worse (*p < 0.001*) than -0.84 ± 0.26 for predicted stables (Supplementary Table 4). Similarly, in the placebo group, predicted progressors had a mean adjusted PACC score of -3.39 ± 0.43 compared to -0.96 ± 0.22 for predicted stables (*p < 0.001*).

**Figure 5.**
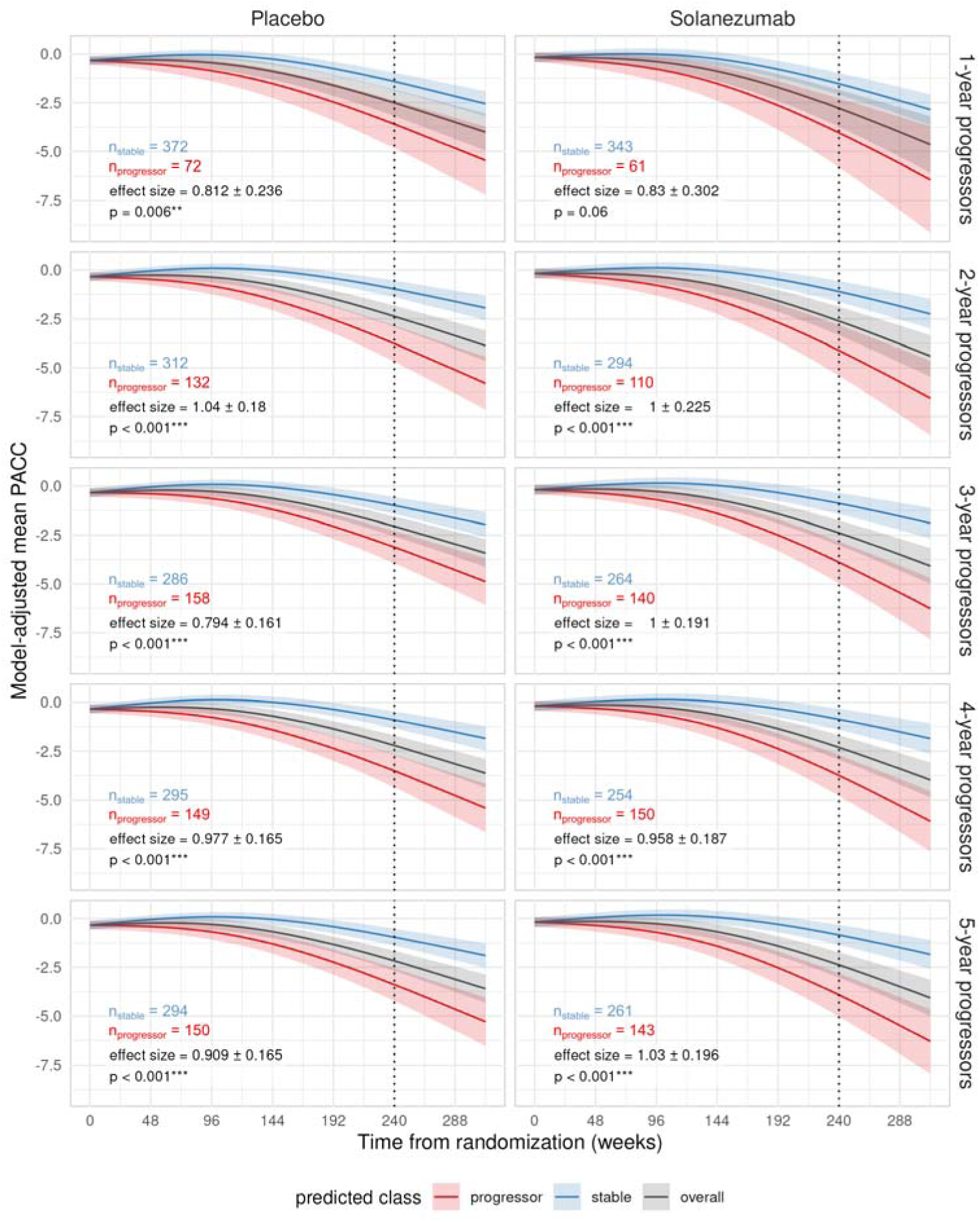
Natural cubic spline models of longitudinal PACC scores of predicted stables and progressors from the A4 study cohort. The model-adjusted PACC trajectory and 95% confidence interval are shown for predicted stables (blue), predicted progressors (red), and the overall cohort (black). The vertical dotted line marks the 240-week follow-up point. The effect size of the contrast between stables and progressors and corresponding p-value are also displayed. Asterisks indicate the significance level (*: p < 0.05, **: p < 0.01, ***: p < 0.001). In all cases except for the 1-year progressors of the Solanezumab arm, significant differences were observed between stables and progressors in the model-adjusted PACC at 240-weeks.

To assess whether cohort enrichment by the trained classifiers improves our ability to detect treatment effects, we tested for significant effects in the primary (PACC) and secondary (FBP cortical SUVR) endpoints after retroactive enrichment. For PACC, we observed no significant difference in the model-adjusted mean PACC score between placebo and Solanezumab arms when the A4 cohort remained unenriched (Table 4), consistent with the conclusions of the original A4 study (Sperling et al., 2023). The difference in model-adjusted mean PACC remained non-significant after enrichment by any model (Table 4), indicating no clear benefit of retroactive enrichment for recovering an underlying treatment effect in the primary outcome.

**Table 4.**
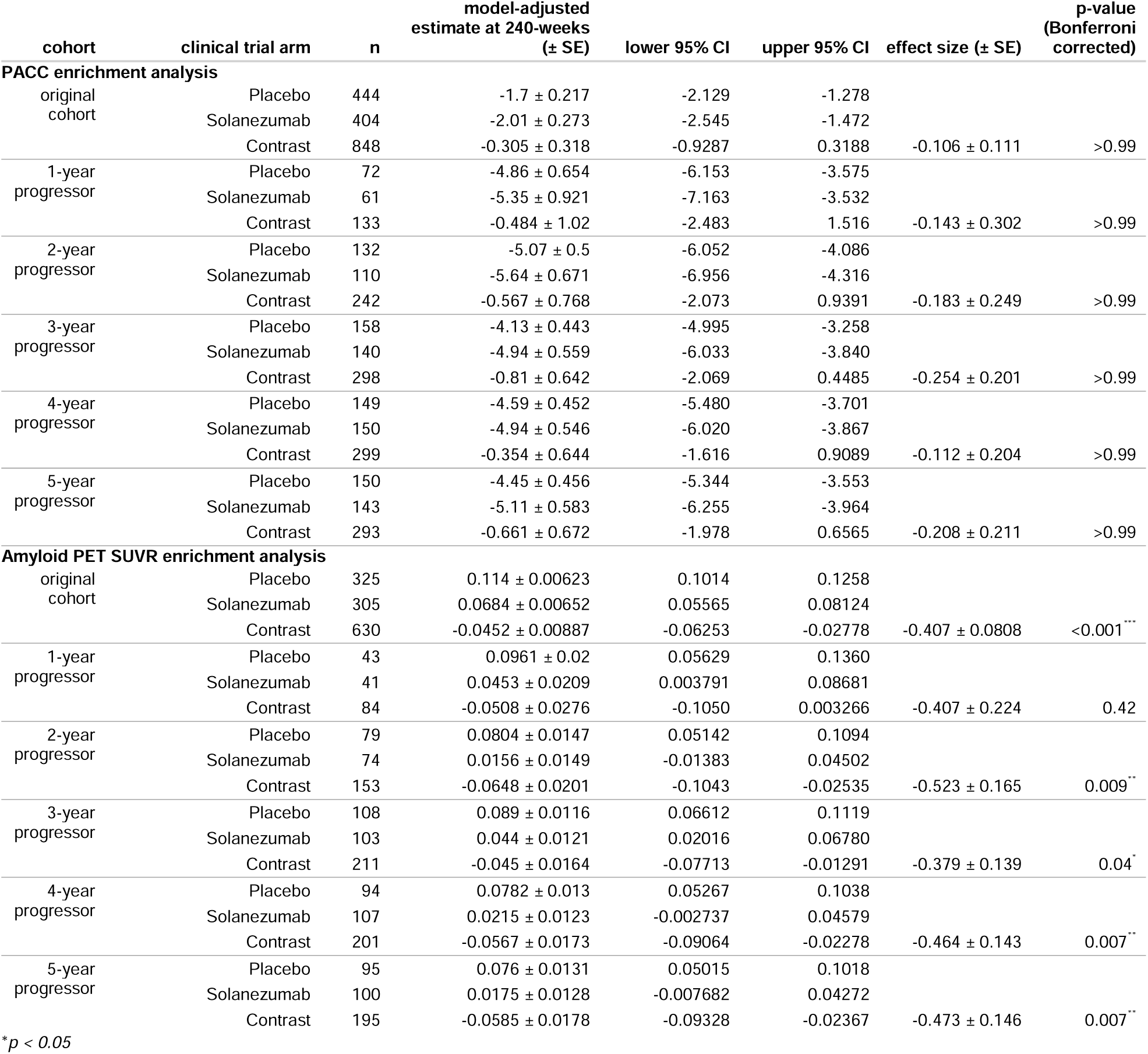
Comparisons of A4 trial outcomes between placebo and Solanezumab arms of the original, unenriched cohort and the ML-enriched cohorts.

For FBP cortical SUVR, a significant difference between trial arms was observed in the mean change in SUVR of the unenriched cohort, with a mean effect size of -0.407 ± 0.08 (*p < 0.001*) (Table 4), suggesting that a reduction in the rate of amyloid accumulation was associated with the treatment. After enrichment, this significant difference was preserved in all but the one-year progressor model. Enrichment by the two, four and five-year models resulted in a modest increase in the mean effect size of the contrast, albeit with an increase in the standard error due to reduced sample sizes. Power analysis revealed that for sample sizes between 20 and 80, an increased power to detect this effect was observed after enrichment by the two, four and five-year models (Figure 6, Supplementary Fig. 5). However, enrichment by the one-year model and, paradoxically, the three-year model showed a decrease in power. This pattern aligned with the results of the ANCOVA in Table 4, wherein mean effect size was increased after enrichment by the two, four and five-year models, but not the one or three-year models.

**Figure 6.**
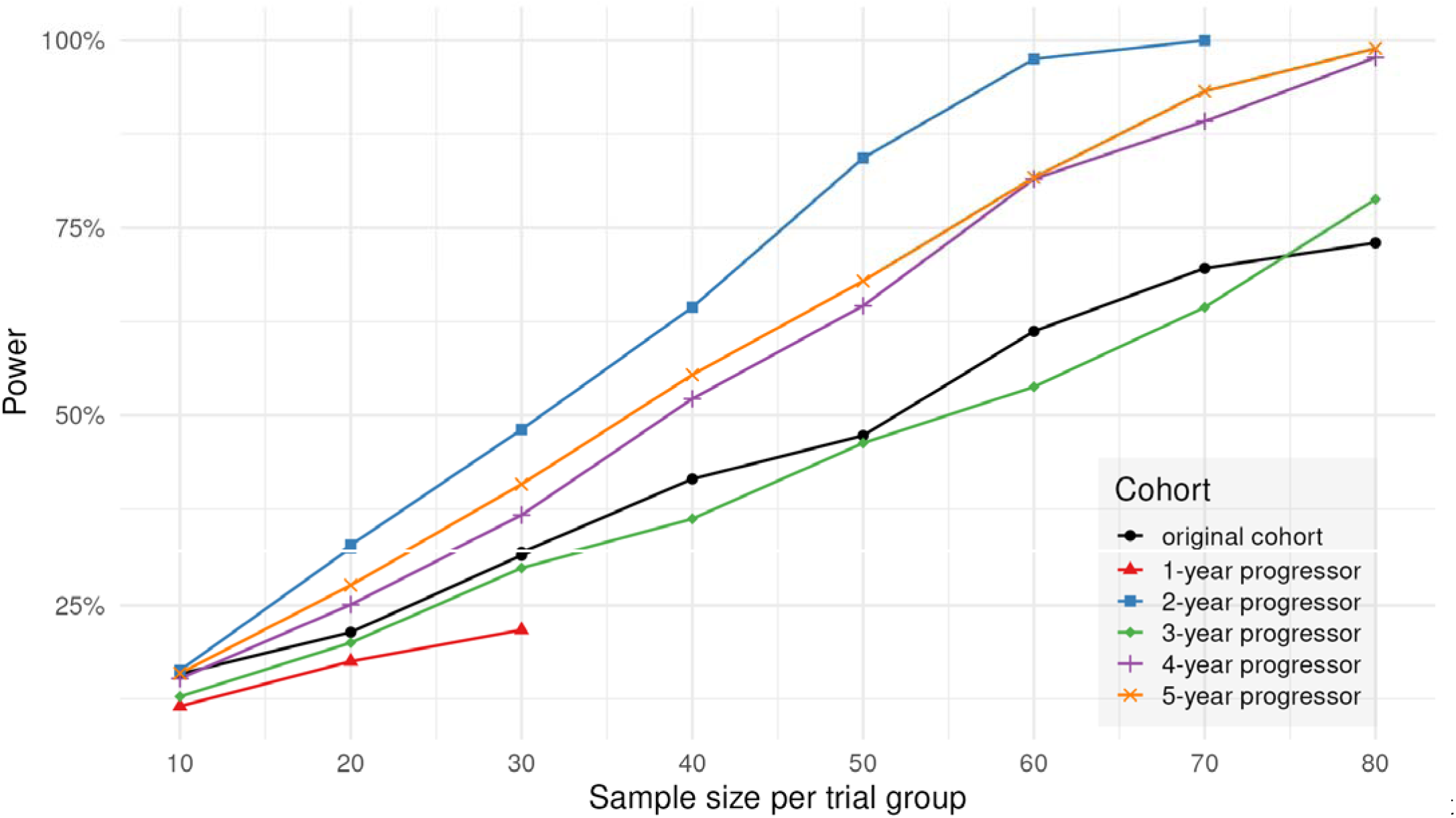
Power to detect differences in change in FBP cortical SUVR between placebo and Solanezumab arms of the A4 study cohort, estimated by bootstrap analysis. Power before (black) and after (colored) retroactive cohort enrichment using SVM classifier predictions are shown. Cohort enrichment by the two-year, four-year and five-year models resulted in increased power at most sample sizes compared to the original, unenriched cohort.

## 4 Discussion

We conducted a rigorous evaluation of machine learning classifiers to predict future cognitive impairment among individuals with preclinical Alzheimer’s disease using imaging features from amyloid PET and MRI. Through leave-one-site-out and leave-one-tracer-out cross-validation, we demonstrated that our models consistently achieved strong generalizability to unseen sites and amyloid PET radiotracers, supporting the viability of these models towards enhancing large, multi-consortia clinical trials for AD. Additionally, we demonstrated that these classifiers may be useful in improving the statistical power of detecting treatment effects using real clinical trial data from the A4 study. The present study represents an important and clinically relevant step towards developing and validating stratification models tailored to the asymptomatic, preclinical AD population. This is of particular importance for future investigations of disease-modifying therapies for AD, given that many are trending towards targeting the disease at an early stage. Furthermore, as amyloid PET is commonly utilized for screening participants into clinical trials (Rafii et al., 2023; Sims et al., 2023; Sperling et al., 2023), our findings suggest that there is added value in quantifying the spatial distribution of amyloid accumulation, beyond a simple dichotomization of global amyloid burden, for stratifying and enriching trial cohorts. In terms of clinical applications, current use cases of amyloid PET include establishing the presence of abnormal pathology before administering anti-amyloid therapy (Cummings et al., 2023), although the use of PET in asymptomatic individuals remains an area of active debate (Leuzy et al., 2025; Rabinovici et al., 2025; Rafii and Aisen, 2023). However, we envision, on the premise that treatments intended for asymptomatic preclinical AD individuals are approved in the future, that amyloid PET coupled with predictive models such as ours would serve as valuable prognostic tools for guiding individualized treatment plans. Such applications are anticipated to expand as mechanisms of reimbursement for amyloid PET are relaxed (“Beta Amyloid Positron Emission Tomography in Dementia and Neurodegenerative Disease,” 2023).

Several prior studies have demonstrated the ability of ML and imaging features extracted from amyloid PET to predict progression to cognitive impairment in CU cohorts with high accuracy (Choi et al., 2024; Dansson et al., 2021; Karaman et al., 2022). The model developed by Karaman *et al*. achieved an ROC-AUC of 0.82 or higher when predicting the future diagnosis of CU individuals from ADNI up to 5-years from baseline. Choi *et al*. reported a mean ROC-AUC of 0.87 when applying their DL-ADprob model to an external CU cohort from HABS to predict future decline at the 4-year follow-up point. In the current study, our classifiers achieved slightly lower performances at comparable follow-up windows. For most leave-out sites, SVM classifiers achieved an out-of-sample ROC-AUC of 0.66 or higher across all times to progression, including ROC-AUC of 0.74 or higher in identifying 4-year progressors or beyond. The notable exception to this was HABS, which scored an ROC-AUC below 0.5. This likely led to high sampling variability that potentially accentuated any existing site-specific biases which the model may not have been robust to. Despite lower comparative performance, we highlight an important distinction of our study compared to previous literature in that we restricted our cohort of interest to CU individuals who showed evidence of abnormal brain amyloidosis, which has been proposed as a core criterion for the biological definition of AD (Jack Jr. et al., 2024). Discrepancies in performance may potentially be attributable to reduced amyloid signal available for differentiating stables from progressors. Additionally, unlike the previously reported classification performance on a HABS CU cohort by (Choi et al., 2024), which was not restricted to be amyloid-positive, our amyloid-positive CU cohort suffered from low sample sizes and extreme imbalances in the stables-to-progressors ratio. Nonetheless, our findings still suggest considerable robustness of SVM classifiers to out-of-sample data for the task of interest, which is crucial for the viability of such models in clinical trial settings for preclinical AD.

In addition to site generalizability, we also showed good generalizability to different amyloid PET radiotracers. Models trained on a single amyloid radiotracer achieved ROC-AUC of 0.72 or greater when applied to the other tracer, even when no explicit tracer harmonization was performed. This, along with previous studies which have shown ML models to accurately predict amyloid positivity (Fan et al., 2024) or future cognitive decline (Choi et al., 2024) without explicit harmonization, may suggest that ML models are capable of finding low-dimensional representations of high-dimensional PET data which are tracer-agnostic. However, we also observed that in the context of a binary classification task, the cutoff thresholds (i.e. the separating hyperplane of the linear SVM) used to determine a binary class decision may not generalize to out-of-sample tracers, as evidenced by high imbalances in the sensitivity and specificity. Converting regional amyloid SUVRs into Centiloids produced models that achieved a much better balance of these metrics, suggesting that explicit harmonization of amyloid PET measurements still holds value for ML models.

While the global amyloid PET signal has been known to be associated with future cognitive decline (Landau et al., 2018; Parent et al., 2023; Small et al., 2012), recent studies have suggested that regional patterns of amyloid accumulation may serve as a more sensitive marker for the risk of cognitive decline, particularly for preclinical AD (Pascoal et al., 2020; Pfeil et al., 2021). By leveraging amyloid PET biomarkers at the regional level, our models were able to discern spatial patterns which discriminate stables from progressors in the preclinical stage. Among the most important amyloid SUVR features were the temporal, superior and middle frontal, and middle occipital cortices. Many of these regions were previously identified as important correlates to increased risk of cognitive decline (Pascoal et al., 2020; Pfeil et al., 2021). Notably, these regions comprise the intermediate stage of amyloid accumulation according to PET staging models (Mattsson et al., 2019). In contrast, our models assigned a lower average feature importance to regions comprising the early accumulation stage, such as the precuneus and orbitofrontal cortices, suggesting that past a global amyloid threshold, the intermediate stage regions become more important for discriminating future cognitive decline.

Additionally, low importances were assigned to subcortical structures such as hippocampus and amygdala, which tend to accumulate amyloid at much later stages of the disease (Hanseeuw et al., 2018; Thal et al., 2002). In terms of MRI features, lateral ventricular volume and the volumes of select subcortical regions such as the hippocampus and amygdala had the highest feature importance. Atrophy in these regions has consistently been identified as robust markers of AD disease progression (Laakso et al., 1995; Mungas et al., 2002; Nestor et al., 2008). Additionally, we found that the importance of amyloid SUVR features increased at a higher rate than volumetric features as the time-to-progression was extended. Our nested model analysis was consistent with this pattern, wherein at higher times-to-progression, the omission of amyloid PET features led to greater decreases in model performance and generalizability compared to either MRI or non-imaging features. These findings may suggest that the influence of amyloid PET on model predictions becomes increasingly important as the cohort includes more early stage progressors. This aligns with current models of AD pathogenesis (Jack Jr. et al., 2024) where amyloid is expected to rise to abnormality well before neurodegeneration occurs, and thus may serve as a better indicator of future cognitive decline in the earlier stages of the disease. Our study notably did not utilize tau PET, which has been shown to closely track longitudinal cognitive decline (Bejanin et al., 2017; Johnson et al., 2016), and may serve as a better predictor of the rate of cognitive decline than either amyloid PET or MRI (Ossenkoppele et al., 2021). Despite this, amyloid PET imaging alone has been shown to provide strong prognostic value for predicting cognitive decline even within amyloid-positive, cognitively normal cohorts (Farrell et al., 2017; Pfeil et al., 2021; van der Kall et al., 2021), a notion which is further supported by our own findings. This is particularly of value for clinical trials for asymptomatic, preclinical AD, where amyloid PET is more often utilized as a screening criterion compared to tau PET.

In the context of cohort recruitment for clinical trials, the clinical presentation of AD may remain highly heterogeneous even after stratification by single risk factors such as global amyloid PET signal or *APOE4* genotype. Alternatively, ML stratification models trained on a combination of multimodal risk factors have been shown to produce more accurate forecasts of longitudinal cognitive decline, leading to greater enhancements in clinical trial operations (Wang et al., 2024). We successfully identified two distinct subtypes within A4 trial participants that exhibited different rates of cognitive decline, where predicted progressors experienced a significantly faster rate of decline compared to stables. However, when applied to cohort enrichment, our models failed to demonstrate a measurable benefit for enhancing the treatment effect in the primary cognitive outcome, PACC. This could be attributed to a true lack of an underlying treatment effect of Solanezumab therapy. Nonetheless, assuming the presence of a ground truth drug effect, ML models may still provide a boost in treatment effect detection as evidenced by previous literature (Birkenbihl et al., 2024; Nallapu et al., 2025; Tam et al., 2022). Interestingly, in terms of the secondary outcome, cortical amyloid deposition, the two, four and five-year classifiers identified enriched cohorts that resulted in an increased power for detecting differences in longitudinal amyloid accumulation. However, this was not the case for either the one-year model, likely due to insufficient sample size selected, or the three-year model, perhaps due to selection of a more heterogeneous cohort that consisted of slightly younger participants with slightly lower cortical SUVRs compared to other enriched cohorts. Given these contradictory findings and the fact that models were not directly trained to predict longitudinal amyloid accumulation, our results should be interpreted with caution. Nonetheless, these results suggest that ML stratification models hold potential to identify trial candidates who are more likely to respond to treatments, and as more clinical trial data become available, future investigations should continue to validate ML models for use within this context.

We note several key limitations which represent areas of future improvement. Firstly, our study did not utilize tau PET, as previously mentioned, and it did not utilize metabolic PET, such as [^18^F]-fluorodeoxyglucose PET, which has also been found to be predictive of cognitive decline in preclinical AD (Mayblyum et al., 2021). For a more comprehensive assessment of AD biomarkers and their prognostic value, future predictive models should integrate these additional modalities. Secondly, due to the strict inclusion criteria employed for cohort selection, the sample sizes in the present study are relatively low compared to other studies employing ML techniques for AD risk prediction. We combatted this by obtaining data from seven independent sites and including PET imaging from two tracers. Additionally, our selected cohort suffered from a high amount of class imbalance, with stable participants heavily outnumbering progressors especially in low times-to-progression. In an effort to reduce the effect of class imbalance, SVM models were optimized with a balanced class weighting, and class-balance invariant metrics such as balanced accuracy and F1 score were reported. Nonetheless, a larger sample of preclinical AD individuals, particularly those who eventually progress to clinical symptoms, is desirable especially for developing more sophisticated ML models such as ones built upon deep learning. Thirdly, given issues of class imbalance, the stable cohort was limited to those who remain CN for at least five years from baseline, representative of a control group with presumably the least amount of pathology. However, a more clinically interpretable model would ideally match the time of stability to the time-to-progression (i.e. an individual predicted not to progress within three years would necessarily remain stable for three years). Lastly, each time-to-progression model was trained independently of the rest, ignoring the constraint that predicted classifications should be consistent across time windows. Future work will explore alternate models such as time-to-event models like the survival SVM or the Mirai model for breast cancer prediction (Yala et al., 2021) which constrain predictions at multiple follow-up times to be temporally consistent.

## 5 Conclusion

We performed an extensive validation of machine learning for predicting future cognitive impairment in cognitively unimpaired individuals at the preclinical stage of AD. Our classifiers maintained strong generalizability across most independent testing sites as well as on unseen amyloid PET radiotracers. We further evaluated our models on the A4 clinical trial cohort and observed potential increases in statistical power to detect a secondary treatment effect, indicating potential utility for cohort enrichment. As emerging investigations of new disease modifying therapies for AD increasingly focus on asymptomatic, preclinical populations, our findings underscore the potential applicability of ML-based patient stratification for recruiting more homogeneous cohorts and improving statistical power for detecting treatment effects for future clinical trials.

## 6 Data availability

Data used in the preparation of this study are openly available and can be requested at https://www.a4studydata.org, https://adni.loni.usc.edu, https://www.mayo.edu/research/centers-programs/alzheimers-disease-research-center/research-activities/mayo-clinic-study-aging/overview, and https://sites.wustl.edu/oasisbrains. HABS data release 1.10, obtained September 2019, was used in the preparation of this article, and can be requested via https://habs.mgh.harvard.edu. A separate application process is involved for accessing PAC. Investigators interested in accessing the data should contact the PAC Coordinating Center at Johns Hopkins University for details.

SVM was implemented in scikit-learn 1.6.0 using python v3.12.8. All statistical analyses were performed in R v4.4.0. Accompanying code will be made publicly available at https://github.com/sotiraslab.

## 7 Acknowledgements

This work was supported by the donors of the Alzheimer’s Disease Research (ADR) program A2021042S, a program of the BrightFocus Foundation. Research reported in this publication was supported by National Institute on Aging of the National Institutes of Health under award number R01AG067103. The content is solely the responsibility of the authors and does not necessarily represent the official views of the National Institutes of Health. The A4 Study was a secondary prevention trial in preclinical Alzheimer’s disease, aiming to slow cognitive decline associated with brain amyloid accumulation in clinically normal older individuals. The A4 Study was funded by a public-private-philanthropic partnership, including funding from the National Institutes of Health-National Institute on Aging, Eli Lilly and Company, Alzheimer’s Association, Accelerating Medicines Partnership, GHR Foundation, an anonymous foundation, and additional private donors, with in-kind support from Avid Radiopharmaceuticals, Cogstate, Albert Einstein College of Medicine and the Foundation for Neurologic Diseases. The companion observational Longitudinal Evaluation of Amyloid Risk and Neurodegeneration (LEARN) Study was funded by the Alzheimer’s Association and GHR Foundation. The A4 and LEARN Studies were led by Dr. Reisa Sperling at Brigham and Women’s Hospital, Harvard Medical School, and Dr. Paul Aisen at the Alzheimer’s Therapeutic Research Institute (ATRI) at the University of Southern California. The A4 and LEARN Studies were coordinated by ATRI at the University of Southern California, and the data are made available under the auspices of Alzheimer’s Clinical Trial Consortium through the Global Research & Imaging Platform (GRIP). The complete A4 Study Team list is available on: https://www.actcinfo.org/a4-study-team-lists/. We would like to acknowledge the dedication of the study participants and their study partners who made the A4 and LEARN Studies possible. Data collection and sharing for this project was funded by the Alzheimer’s Disease Neuroimaging Initiative (ADNI) (National Institutes of Health Grant U01 AG024904) and DOD ADNI (Department of Defense award number W81XWH-12-2-0012). ADNI is funded by the National Institute on Aging, the National Institute of Biomedical Imaging and Bioengineering, and through generous contributions from the following: AbbVie, Alzheimer’s Association; Alzheimer’s Drug Discovery Foundation; Araclon Biotech; BioClinica, Inc.; Biogen; Bristol-Myers Squibb Company; CereSpir, Inc.; Cogstate; Eisai Inc.; Elan Pharmaceuticals, Inc.; Eli Lilly and Company; EuroImmun; F. Hoffmann-La Roche Ltd and its affiliated company Genentech, Inc.; Fujirebio; GE Healthcare; IXICO Ltd.; Janssen Alzheimer Immunotherapy Research & Development, LLC.; Johnson & Johnson Pharmaceutical Research & Development LLC.; Lumosity; Lundbeck; Merck & Co., Inc.; Meso Scale Diagnostics, LLC.; NeuroRx Research; Neurotrack Technologies; Novartis Pharmaceuticals Corporation; Pfizer Inc.; Piramal Imaging; Servier; Takeda Pharmaceutical Company; and Transition Therapeutics. The Canadian Institutes of Health Research is providing funds to support ADNI clinical sites in Canada. Private sector contributions are facilitated by the Foundation for the National Institutes of Health (www.fnih.org). The grantee organization is the Northern California Institute for Research and Education, and the study is coordinated by the Alzheimer’s Therapeutic Research Institute at the University of Southern California. ADNI data are disseminated by the Laboratory for Neuro Imaging at the University of Southern California. Part of the data used in the preparation of this article were obtained from the Harvard Aging Brain Study (HABS - P01AG036694; https://nam10.safelinks.protection.outlook.com/?url=https%3A%2F%2Fhabs.mgh.harvard.edu%2F&data=05%7C02%7Ctom.earnest%40wustl.edu%7C70bcf8c15ffb44a9848408dccc4c749f%7C4ccca3b571cd4e6d974b4d9beb96c6d6%7C0%7C0%7C638609875978514619%7CUnknown%7CTWFpbGZsb3d8eyJWIjoiMC4wLjAwMDAiLCJQIjoiV2luMzIiLCJBTiI6Ik1haWwiLCJXVCI6Mn0%3D%7C0%7C%7C%7C&sdata=uZIOX1693oz1jyd49mvRaqaaIHC%2BXKDKPHUdotwp%2B8I%3D&reserved=0). The HABS study was launched in 2010, funded by the National Institute on Aging. and is led by principal investigators Reisa A. Sperling MD and Keith A. Johnson MD at Massachusetts General Hospital/Harvard Medical School in Boston, MA. Part of the data contained in this analysis were obtained under one of the following research grants from the National Institutes of Health to the Mayo Clinic Study of Aging (U01 AG06786, Ronald Petersen, PI) or the Mayo Alzheimer’s Disease Research Center (P50 AG16574, Ronald Petersen, PI). Part of the data were provided by OASIS-3: Longitudinal Multimodal Neuroimaging (Principal Investigators: T. Benzinger, D. Marcus, J. Morris). OASIS-3 was supported by the following funding sources: NIH P30 AG066444, P30 NS09857781, P01 AG026276, P01 AG003991, R01 AG043434, UL1 TR000448, R01 EB009352. AV-45 doses were provided by Avid Radiopharmaceuticals, a wholly owned subsidiary of Eli Lilly. Avid Radiopharmaceuticals, Inc., a wholly owned subsidiary of Eli Lilly and company, enabled use of the 18F-florbetapir tracer, but did not provide direct funding and was not involved in data analysis or interpretation. The Preclinical AD Consortium is supported by the National Institutes of Health (NIH), USA [grant number RF1-AG059869]. The individual studies in the consortium are funded, in part, by the following grants: U19-AG033655, P01-AG026276, RF1-AG027161 and the Australian Commonwealth Scientific Industrial Research Organization (CSIRO); as well as the National Institute on Aging Intramural Research Program.

## 8 CRediT authorship contribution statement

**Braden Yang**: Conceptualization, Investigation, Formal analysis, Methodology, Software, Visualization, Writing - original draft. **Tom Earnest**: Methodology, Software, Data curation, Writing - review & editing. **Murat Bilgel**: Methodology, Software, Data curation, Writing - review & editing. **Marilyn S. Albert**: Data curation. **Sterling C. Johnson**: Data curation. **Christos Davatzikos**: Data curation. **Guray Erus**: Data curation. **Colin L. Masters**: Data curation. **Susan M. Resnick**: Data curation, Writing - review & editing. **Michael I. Miller**: Data curation. **Arnold Bakker**: Data curation. **John C. Morris**: Data curation, Writing - review & editing. **Tammie L.S. Benzinger**: Conceptualization, Supervision, Data curation, Writing - review & editing. **Brian A. Gordon**: Conceptualization, Supervision, Writing - review & editing. **Aristeidis Sotiras**: Conceptualization, Supervision, Project administration, Funding acquisition, Writing - review & editing.

