## Supplementary material for "Predicting future cognitive impairment in preclinical Alzheimer’s disease using amyloid PET and MRI: a multisite machine learning study"

**Supplementary Methods**

The following image acquisition and processing procedures were performed independently by each data collecting site. Note that these were used solely for the purposes of determining amyloid-positivity for selection of our study cohort. Amyloid PET and MRI features used for training SVM classifiers were obtained using a separate pipeline detailed in the main text.

**A4**

Participants received [^18^F]-florbetapir amyloid PET imaging, with data acquired 50-70 minutes post-injection. Semiautomated quantitative analysis of PET images was performed as previously described (Clark et al., 2012; Joshi et al., 2015). Mean cortical standard uptake value ratios (SUVR) with whole cerebellum as reference region was computed from the mean signal of 6 cortical regions: medial orbital frontal, anterior cingulate, temporal, precuneus, parietal, and posterior cingulate. Scans were labeled as amyloid-positive if their mean cortical SUVR was 1.15 or higher (Sperling et al., 2020).

**ADNI**

All participants selected from the ADNI database for this study were imaged using [^18^F]-florbetapir. Participants received a 370 MBq injection of [^18^F]-florbetapir, followed by acquisition of four 5-minute frames from 50-70 minutes post-injection. Processing of PET images is detailed elsewhere (Landau et al., 2025). Briefly, FBP frames were coregistered, averaged, standardized and resampled to uniform voxel resolution. Images were then smoothed to 6mm full width at half maximum (FWHM). For older scans that were fixed at 8mm FWHM, linear regression equations were developed to transform SUVRs from 8mm scans into equivalent 6mm versions. Accompanying MRI scans were segmented and parcellated with FreeSurfer version 7.1.1, and a cortical summary region was constructed using frontal, anterior cingulate, posterior cingulate, lateral parietal, and lateral temporal regions. Mean cortical SUVRs were computed using whole cerebellum as reference region. Scans were labeled as amyloid-positive if their mean cortical SUVR was 1.11 or higher.

**HABS**

A complete description of the HABS dataset can be found in (Dagley et al., 2017). All amyloid PET scans were acquired at Massachusetts General Hospital in Boston, MA, and all MRI scans were acquired at MGH-Martinos Center for Biomedical Imaging in Charlestown, MA. Participants were imaged using [^11^C]-Pittsburgh compound-B. Global SUVRs were computed in a cortical composite region using cerebellar gray matter as reference region. Scans were labeled as amyloid-positive if their global SUVR was 1.28 or higher.

**MCSA**

All participants were recruited from the Mayo Alzheimer’s Disease Research Center or Alzheimer’s Disease Patient Registry. Participants received a mean 628 MBq (385-723 MBq) injection of [^11^C]-Pittsburgh compound-B. 5-minute dynamic PET frames from 40-60 minutes post-injection were acquired. Cortical global SUVR and Centiloid values were computed with cerebellar gray matter as reference region. Scans were labeled as amyloid-positive if their Centiloid was 21.5 or higher.

**OASIS-3**

A complete description of the OASIS-3 dataset can be found in (LaMontagne et al., 2019). All amyloid PET and MRI scans were acquired at Washington University in St. Louis. For PiB PET, participants received a bolus injection of 6–20 mCi of PiB and a 60-minute dynamic scan was acquired. For FBP PET, participants received a bolus injection of 10 mCi of FBP, and either a 70-min dynamic scan was acquired, or a 20-min dynamic scan was acquired at 50-min post-injection.

T1-weighted MRI scans were segmented and parcellated into regions-of-interest using FreeSurfer 5.0 or 5.1 for 1.5T scans or FreeSurfer 5.3 for 3T scans. To determine amyloid-positivity, all PET images were processed using the PET Unified Pipeline (Su et al., 2013). Briefly, PET frames were selected within a post-injection time window (30 to 60 minutes for PiB, 50 to 70 minutes for FBP), smoothed to 8-mm spatial resolution, corrected for inter-frame motion, and coregistered to the T1 MRI scan acquired closest in time using a vector-gradient algorithm. Global cortical amyloid deposition was quantified by taking the average signal across the lateral and medial orbitofrontal, middle and superior temporal, superior frontal, rostral middle frontal, and precuneus FreeSurfer regions-of-interest from both hemispheres. The average of the left and right cerebellar cortex was used to normalize the global cortical amyloid signal to an SUVR. Scans were labeled as amyloid-positive if their global cortical SUVR was 1.31 or higher for PiB and 1.24 or higher for FBP (Su et al., 2019). We utilized the non-partial volume corrected SUVRs to determine amyloid-positivity.

**Preclinical Alzheimer’s Consortium**

Using T1-weighted MRI scans, we computed anatomical labels and regional brain volumes using Multi-atlas region Segmentation using Ensembles of registration algorithms and parameters (MUSE) (Doshi et al., 2016), and intracranial masks were computed using a deep learning approach (Doshi et al., 2019). In the BLSA, scanner-specific atlases were used to harmonize MUSE processing across 1.5 T and 3 T scanners (Erus et al., 2018).

11C-Pittsburgh compound B (PiB) PET data provided by AIBL included summed images (not motion-corrected) within the 50-70 min post-injection time window. The remaining four studies (BIOCARD, BLSA, OASIS3, and WRAP) in the Preclinical Alzheimer’s Disease Consortium had dynamic PiB PET scans obtained for at least 60 min immediately following an intravenous bolusinjection of radiotracer. Each PET scan was matched with the MRI closest in time. For inclusion in these analyses, we required a T1-weighted MRI within 5 years of PET.

For the dynamic acquisitions, we aligned each PET time frame to the mean of the frames in the first 2 min to correct for motion using the Realign algorithm in SPM (https://www.fil.ion.ucl.ac.uk/spm/software/spm12/, SPM12-r7219) (Ashburner and Friston, 1997). Participants who had a translation > 2 cm or rotation > 0.1 rad about the x, y, or z-axis at any time frame were excluded. We registered the smoothed (with a 4 mm FWHM Gaussian filter) average of the 50-60 min or 50-70 min of realigned scans onto the corresponding inhomogeneity-corrected and skull-stripped T1-weighted MRI scan using rigid registration with mutual information using the FLIRT algorithm (https://fsl.fmrib.ox.ac.uk/fsl, version 6.0.3) (Jenkinson et al., 2012, 2002). During coregistration optimization, calculation of mutual information was restricted to PET voxels with at least one count to account for field of view differences between the scans. We examined registration parameters of the coregistered PET and MRI images to identify outliers for further visual inspection. Any misregistrations identified upon visual inspection were excluded. Using the coregistration result, we transformed the MUSE label image from MRI to PET space. We inspected time activity curves in cortical regions to detect and exclude any irregular scans (i.e., scans whose acquisitions started late, interrupted scans, or scans where cerebral cortical to cerebellar cortical signal ratio markedly decreased over time). MUSE anatomical labels defined in MRI space were transformed into PET space using the coregistration result and masked to voxels within the PET field of view. We computed standardized uptake value ratio (SUVR) images by dividing the PET image by the mean within the cerebellar gray matter. We calculated mean cortical amyloid burden as the average of the SUVR values in cingulate, frontal, parietal (including precuneus), lateral temporal, and lateral occipital cortical regions, excluding the sensorimotor strip. PET image processing steps were streamlined using nipype (<https://nipype.readthedocs.io/>, version 1.6.0) (Gorgolewski et al., 2011) in Python 3.8.6. PiB PET scans in AIBL were provided as summed images within 50-70 min post-injection (where at least 15 min of data are available). To be compatible with the AIBL data, we calculated SUVR within 50-70 min in BIOCARD, BLSA, and WRAP for scans with at least 15 min of data within this time window. We could not compute 50-70 min SUVR in OASIS3 (except for one scan) since acquisitions do not go beyond min 60. Instead, we computed 50-60 min SUVR in OASIS3 and imputed 50-70 min SUVR. To do this, we first estimated the intercept and regression coefficient of 50-60 min SUVR in a linear regression model with 50-70 min SUVR as the outcome using pooled data from BIOCARD, BLSA, and WRAP. We then used these estimates to predict 50-70 min SUVR in OASIS3 using 50-60 min SUVR. For some OASIS3 scans, 50-60 min SUVRs were not available due to insufficient data within this time interval, but DVR and R1 data were available (see PiB DVR report for detailed descriptions of DVR and R1 calculations). For these scans, we predicted 50-70 min SUVR using a linear regression model including intercept, DVR, and R1 trained on pooled data from BIOCARD, BLSA, and WRAP. To impute 50-70 min SUVR, we fitted separate linear regression models for each ROI.

We categorized each individual as amyloid –/+ based on a mean cortical SUVR threshold of 1.237, which was derived from a Gaussian mixture model fitted to baseline mean cortical SUVR values pooled across the four sites using the mixtools package (version 1.2.0) (Benaglia et al., 2010) in R version 4.0.3.

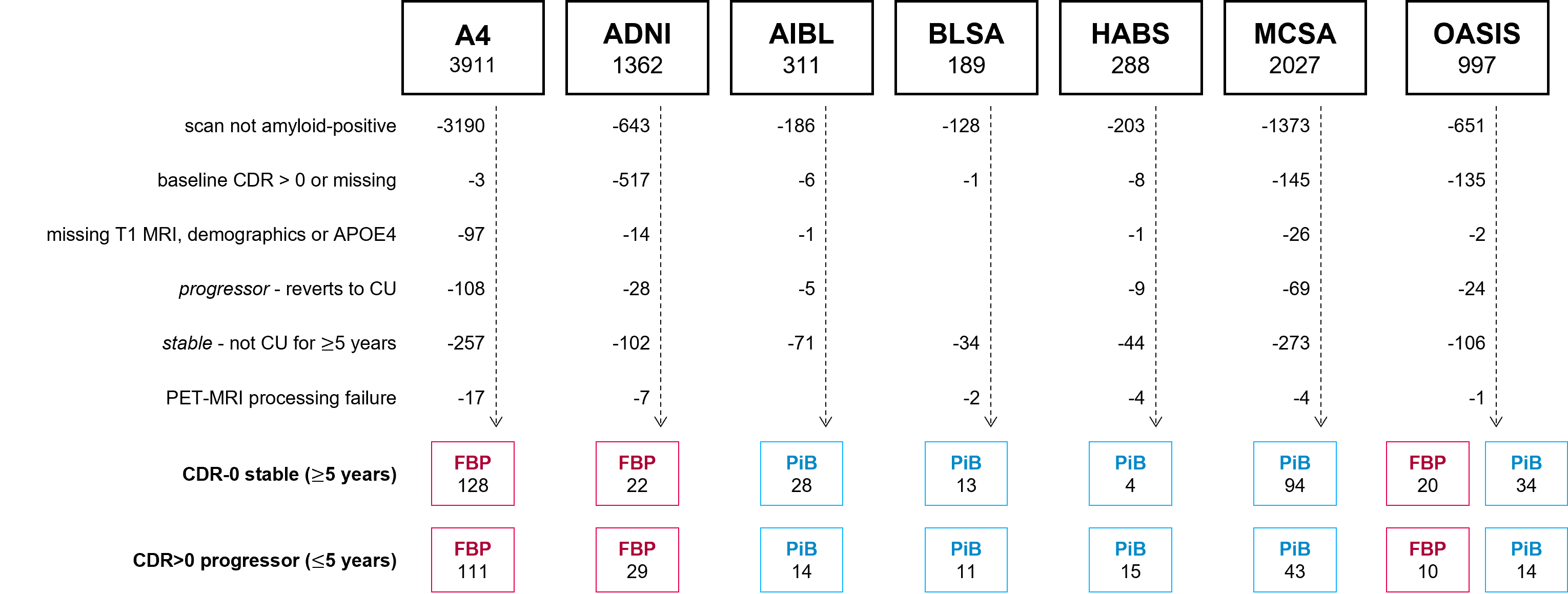

**Supplementary Figure 1.** Subject selection flowchart.

**Supplementary Table 1.** List of MUSE regions used as input features for SVM classifiers.

| **ROI name** | **ROI type** | **amyloid PET SUVR** | **volumetric** |
| --- | --- | --- | --- |
| Right Accumbens Area | subcortical | yes | yes |
| Left Accumbens Area | subcortical | yes | yes |
| Right Amygdala | subcortical | yes | yes |
| Left Amygdala | subcortical | yes | yes |
| Right Caudate | subcortical | yes | yes |
| Left Caudate | subcortical | yes | yes |
| Right Hippocampus | subcortical | yes | yes |
| Left Hippocampus | subcortical | yes | yes |
| Right Pallidum | subcortical | yes | yes |
| Left Pallidum | subcortical | yes | yes |
| Right Putamen | subcortical | yes | yes |
| Left Putamen | subcortical | yes | yes |
| Right Thalamus Proper | subcortical | yes | yes |
| Left Thalamus Proper | subcortical | yes | yes |
| Left Basal Forebrain | subcortical | yes | yes |
| Right Basal Forebrain | subcortical | yes | yes |
| Right ACgG anterior cingulate gyrus | cortical | yes | yes |
| Left ACgG anterior cingulate gyrus | cortical | yes | yes |
| Right AIns anterior insula | cortical | yes | yes |
| Left AIns anterior insula | cortical | yes | yes |
| Right AOrG anterior orbital gyrus | cortical | yes | yes |
| Left AOrG anterior orbital gyrus | cortical | yes | Yes |
| Right AnG angular gyrus | cortical | yes | Yes |
| Left AnG angular gyrus | cortical | yes | Yes |
| Right Calc calcarine cortex | cortical | yes | yes |
| Left Calc calcarine cortex | cortical | yes | yes |
| Right CO central operculum | cortical | yes | yes |
| Left CO central operculum | cortical | yes | yes |
| Right Cun cuneus | cortical | yes | yes |
| Left Cun cuneus | cortical | yes | yes |
| Right Ent entorhinal area | cortical | yes | yes |
| Left Ent entorhinal area | cortical | yes | yes |
| Right FO frontal operculum | cortical | yes | yes |
| Left FO frontal operculum | cortical | yes | yes |
| Right FRP frontal pole | cortical | yes | yes |
| Left FRP frontal pole | cortical | yes | yes |
| Right FuG fusiform gyrus | cortical | yes | yes |
| Left FuG fusiform gyrus | cortical | yes | yes |
| Right GRe gyrus rectus | cortical | yes | yes |
| Left GRe gyrus rectus | cortical | yes | yes |
| Right IOG inferior occipital gyrus | cortical | yes | yes |
| Left IOG inferior occipital gyrus | cortical | yes | yes |
| Right ITG inferior temporal gyrus | cortical | yes | yes |
| Left ITG inferior temporal gyrus | cortical | yes | yes |
| Right LiG lingual gyrus | cortical | yes | yes |
| Left LiG lingual gyrus | cortical | yes | yes |
| Right LOrG lateral orbital gyrus | cortical | yes | yes |
| Left LOrG lateral orbital gyrus | cortical | yes | yes |
| Right MCgG middle cingulate gyrus | cortical | yes | yes |
| Left MCgG middle cingulate gyrus | cortical | yes | yes |
| Right MFC medial frontal cortex | cortical | yes | yes |
| Left MFC medial frontal cortex | cortical | yes | yes |
| Right MFG middle frontal gyrus | cortical | yes | yes |
| Left MFG middle frontal gyrus | cortical | yes | yes |
| Right MOG middle occipital gyrus | cortical | yes | yes |
| Left MOG middle occipital gyrus | cortical | yes | yes |
| Right MOrG medial orbital gyrus | cortical | yes | yes |
| Left MOrG medial orbital gyrus | cortical | yes | yes |
| Right MPoG postcentral gyrus medial segment | cortical | yes | yes |
| Left MPoG postcentral gyrus medial segment | cortical | yes | yes |
| Right MPrG precentral gyrus medial segment | cortical | yes | yes |
| Left MPrG precentral gyrus medial segment | cortical | yes | yes |
| Right MSFG superior frontal gyrus medial segment | cortical | yes | yes |
| Left MSFG superior frontal gyrus medial segment | cortical | yes | yes |
| Right MTG middle temporal gyrus | cortical | yes | yes |
| Left MTG middle temporal gyrus | cortical | yes | yes |
| Right OCP occipital pole | cortical | yes | yes |
| Left OCP occipital pole | cortical | yes | yes |
| Right OFuG occipital fusiform gyrus | cortical | yes | yes |
| Left OFuG occipital fusiform gyrus | cortical | yes | yes |
| Right OpIFG opercular part of the inferior frontal gyrus | cortical | yes | yes |
| Left OpIFG opercular part of the inferior frontal gyrus | cortical | yes | yes |
| Right OrIFG orbital part of the inferior frontal gyrus | cortical | yes | yes |
| Left OrIFG orbital part of the inferior frontal gyrus | cortical | yes | yes |
| Right PCgG posterior cingulate gyrus | cortical | yes | yes |
| Left PCgG posterior cingulate gyrus | cortical | yes | yes |
| Right PCu precuneus | cortical | yes | yes |
| Left PCu precuneus | cortical | yes | yes |
| Right PHG parahippocampal gyrus | cortical | yes | yes |
| Left PHG parahippocampal gyrus | cortical | yes | yes |
| Right PIns posterior insula | cortical | yes | yes |
| Left PIns posterior insula | cortical | yes | yes |
| Right PO parietal operculum | cortical | yes | yes |
| Left PO parietal operculum | cortical | yes | yes |
| Right PoG postcentral gyrus | cortical | yes | yes |
| Left PoG postcentral gyrus | cortical | yes | yes |
| Right POrG posterior orbital gyrus | cortical | yes | yes |
| Left POrG posterior orbital gyrus | cortical | yes | yes |
| Right PP planum polare | cortical | yes | yes |
| Left PP planum polare | cortical | yes | yes |
| Right PrG precentral gyrus | cortical | yes | yes |
| Left PrG precentral gyrus | cortical | yes | yes |
| Right PT planum temporale | cortical | yes | yes |
| Left PT planum temporale | cortical | yes | yes |
| Right SCA subcallosal area | cortical | yes | yes |
| Left SCA subcallosal area | cortical | yes | yes |
| Right SFG superior frontal gyrus | cortical | yes | yes |
| Left SFG superior frontal gyrus | cortical | yes | yes |
| Right SMC supplementary motor cortex | cortical | yes | yes |
| Left SMC supplementary motor cortex | cortical | yes | yes |
| Right SMG supramarginal gyrus | cortical | yes | yes |
| Left SMG supramarginal gyrus | cortical | yes | yes |
| Right SOG superior occipital gyrus | cortical | yes | yes |
| Left SOG superior occipital gyrus | cortical | yes | yes |
| Right SPL superior parietal lobule | cortical | yes | yes |
| Left SPL superior parietal lobule | cortical | yes | yes |
| Right STG superior temporal gyrus | cortical | yes | yes |
| Left STG superior temporal gyrus | cortical | yes | yes |
| Right TMP temporal pole | cortical | yes | yes |
| Left TMP temporal pole | cortical | yes | yes |
| Right TrIFG triangular part of the inferior frontal gyrus | cortical | yes | yes |
| Left TrIFG triangular part of the inferior frontal gyrus | cortical | yes | yes |
| Right TTG transverse temporal gyrus | cortical | yes | yes |
| Left TTG transverse temporal gyrus | cortical | yes | yes |
| 3rd Ventricle | ventricle | no | yes |
| 4th Ventricle | ventricle | no | yes |
| Right Inf Lat Vent | ventricle | no | yes |
| Left Inf Lat Vent | ventricle | no | yes |
| Right Lateral Ventricle | ventricle | no | yes |
| Left Lateral Ventricle | ventricle | no | yes |

**Supplementary Table 2.** Sensitivity analysis results of SVM classifiers trained on unharmonized (*base*) and harmonized (*Centiloid*) regional amyloid PET SUVRs. For base experiments, the raw evaluation metric was reported, whereas for Centiloid experiments, the difference in metric relative to base was reported.

|  |  |  | **leave-out group** | | | | | | | |
| --- | --- | --- | --- | --- | --- | --- | --- | --- | --- | --- |
|  | **progression time-horizon (yrs)** | **experiment** | **A4** | **ADNI** | **MCSA** | **OASIS** | **aggregated (site)** | **FBP** | **PIB** | **aggregated (tracer)** |
| ROC AUC | 1 | base | 0.8391 | 0.7803 | 0.8404 | 0.9339 | - | 0.7710 | 0.9082 | - |
|  |  | Centiloid | -0.0453 | +0.0152 | -0.0160 | +0.0026 | - | -0.0681 | -0.0459 | - |
|  | 2 | base | 0.7104 | 0.8182 | 0.8304 | 0.9630 | - | 0.7398 | 0.8462 | - |
|  |  | Centiloid | +0.0151 | +0.0265 | +0.0038 | -0.0021 | - | -0.0115 | +0.0153 | - |
|  | 3 | base | 0.7421 | 0.8455 | 0.7531 | 0.8725 | - | 0.7713 | 0.7691 | - |
|  |  | Centiloid | -0.0434 | -0.0023 | -0.0063 | -0.0033 | - | -0.0038 | -0.0227 | - |
|  | 4 | base | 0.7590 | 0.8011 | 0.7670 | 0.8519 | - | 0.7738 | 0.7876 | - |
|  |  | Centiloid | -0.0135 | +0.0057 | -0.0058 | -0.0107 | - | -0.0035 | +0.0044 | - |
|  | 5 | base | 0.7553 | 0.7821 | 0.7199 | 0.8488 | - | 0.7699 | 0.6945 | - |
|  |  | Centiloid | -0.0127 | +0.0016 | -0.0106 | -0.0394 | - | -0.0065 | +0.0964 | - |
| Accuracy | 1 | base | 0.8421 | 0.7500 | 0.6735 | 0.9016 | 0.7937 | 0.9239 | 0.6985 | 0.8281 |
|  |  | Centiloid | -0.1278 | +0.0357 | 0.0000 | -0.0164 | -0.0531 | -0.0815 | -0.0074 | -0.0500 |
|  | 2 | base | 0.7824 | 0.7059 | 0.6126 | 0.9206 | 0.7487 | 0.7719 | 0.6067 | 0.7063 |
|  |  | Centiloid | -0.0882 | +0.0294 | +0.1892 | +0.0159 | +0.0212 | -0.0044 | +0.1000 | +0.0370 |
|  | 3 | base | 0.7438 | 0.8095 | 0.6484 | 0.8732 | 0.7432 | 0.7216 | 0.6082 | 0.6779 |
|  |  | Centiloid | -0.0936 | -0.0476 | +0.0859 | -0.0141 | -0.0248 | +0.0037 | -0.0117 | -0.0023 |
|  | 4 | base | 0.7309 | 0.7826 | 0.6176 | 0.8219 | 0.7176 | 0.7037 | 0.6464 | 0.6820 |
|  |  | Centiloid | -0.0493 | -0.0217 | +0.0882 | 0.0000 | 0.0000 | +0.0168 | +0.0718 | +0.0377 |
|  | 5 | base | 0.7113 | 0.8039 | 0.5839 | 0.7821 | 0.6970 | 0.6719 | 0.5622 | 0.6317 |
|  |  | Centiloid | -0.0669 | -0.0784 | -0.0146 | 0.0000 | -0.0436 | +0.0406 | +0.1405 | +0.0772 |
| Balanced Accuracy | 1 | base | 0.7258 | 0.5379 | 0.7101 | 0.6958 | 0.6367 | 0.5983 | 0.7812 | 0.6551 |
|  |  | Centiloid | -0.1625 | +0.0833 | 0.0000 | -0.0714 | -0.0496 | +0.0214 | -0.0039 | +0.0153 |
|  | 2 | base | 0.6875 | 0.7159 | 0.6990 | 0.9074 | 0.7126 | 0.5801 | 0.7131 | 0.5852 |
|  |  | Centiloid | -0.0106 | +0.0417 | +0.0153 | +0.0093 | +0.0271 | +0.0255 | +0.0398 | +0.0418 |
|  | 3 | base | 0.7003 | 0.8114 | 0.6762 | 0.8361 | 0.7196 | 0.6559 | 0.6997 | 0.6413 |
|  |  | Centiloid | -0.0521 | -0.0477 | -0.0072 | -0.0496 | -0.0219 | +0.0508 | -0.0155 | +0.0402 |
|  | 4 | base | 0.7127 | 0.7803 | 0.6641 | 0.7602 | 0.7086 | 0.6675 | 0.6837 | 0.6504 |
|  |  | Centiloid | -0.0172 | -0.0208 | +0.0177 | -0.0341 | +0.0099 | +0.0456 | +0.0066 | +0.0555 |
|  | 5 | base | 0.7047 | 0.7947 | 0.6400 | 0.7269 | 0.6946 | 0.6543 | 0.6057 | 0.6068 |
|  |  | Centiloid | -0.0505 | -0.0690 | -0.0296 | +0.0116 | -0.0266 | +0.0598 | +0.0821 | +0.1031 |
| F1 Score | 1 | base | 0.2222 | 0.2222 | 0.1579 | 0.5000 | 0.2326 | 0.3000 | 0.2545 | 0.2667 |
|  |  | Centiloid | -0.1270 | +0.1778 | 0.0000 | -0.1364 | -0.0543 | -0.0436 | -0.0045 | -0.0140 |
|  | 2 | base | 0.5316 | 0.6429 | 0.3944 | 0.7619 | 0.5226 | 0.2973 | 0.3918 | 0.3509 |
|  |  | Centiloid | -0.0222 | +0.0468 | +0.0818 | +0.0381 | +0.0358 | +0.0792 | +0.0582 | +0.0612 |
|  | 3 | base | 0.6061 | 0.8095 | 0.5263 | 0.7429 | 0.6250 | 0.5128 | 0.5315 | 0.5217 |
|  |  | Centiloid | -0.0312 | -0.0476 | -0.0120 | -0.0554 | -0.0269 | +0.1213 | -0.0140 | +0.0645 |
|  | 4 | base | 0.6512 | 0.8000 | 0.5593 | 0.6486 | 0.6419 | 0.5464 | 0.5616 | 0.5529 |
|  |  | Centiloid | +0.0276 | -0.0245 | +0.0059 | -0.0426 | +0.0163 | +0.1229 | +0.0025 | +0.0829 |
|  | 5 | base | 0.6634 | 0.8333 | 0.5440 | 0.6222 | 0.6483 | 0.5161 | 0.5031 | 0.5105 |
|  |  | Centiloid | +0.0109 | -0.0833 | -0.0316 | +0.0161 | -0.0091 | +0.1909 | +0.0706 | +0.1576 |
| Sensitivity | 1 | base | 0.6000 | 0.1667 | 0.7500 | 0.4286 | 0.4545 | 0.2143 | 0.8750 | 0.4545 |
|  |  | Centiloid | -0.2000 | +0.1667 | 0.0000 | -0.1429 | -0.0455 | +0.1429 | 0.0000 | +0.0909 |
|  | 2 | base | 0.5000 | 0.7500 | 0.8235 | 0.8889 | 0.6500 | 0.1897 | 0.8636 | 0.3750 |
|  |  | Centiloid | +0.1429 | +0.0833 | -0.2353 | 0.0000 | +0.0375 | +0.0862 | -0.0455 | +0.0500 |
|  | 3 | base | 0.5333 | 0.8500 | 0.7353 | 0.7647 | 0.6507 | 0.3883 | 0.8837 | 0.5342 |
|  |  | Centiloid | +0.1067 | -0.0500 | -0.2059 | -0.1176 | -0.0137 | +0.2427 | -0.0233 | +0.1644 |
|  | 4 | base | 0.5895 | 0.8333 | 0.7857 | 0.6316 | 0.6722 | 0.4173 | 0.7736 | 0.5222 |
|  |  | Centiloid | +0.2000 | -0.0417 | -0.1667 | -0.1053 | +0.0500 | +0.2441 | -0.1509 | +0.1278 |
|  | 5 | base | 0.6126 | 0.8621 | 0.7907 | 0.5833 | 0.6812 | 0.3733 | 0.7193 | 0.4686 |
|  |  | Centiloid | +0.1802 | -0.1379 | -0.0698 | +0.0417 | +0.0676 | +0.3667 | -0.0702 | +0.2464 |
| Specificity | 1 | base | 0.8516 | 0.9091 | 0.6702 | 0.9630 | 0.8188 | 0.9824 | 0.6875 | 0.8557 |
|  |  | Centiloid | -0.1250 | 0.0000 | 0.0000 | 0.0000 | -0.0537 | -0.1000 | -0.0078 | -0.0604 |
|  | 2 | base | 0.8750 | 0.6818 | 0.5745 | 0.9259 | 0.7752 | 0.9706 | 0.5625 | 0.7953 |
|  |  | Centiloid | -0.1641 | 0.0000 | +0.2660 | +0.0185 | +0.0168 | -0.0353 | +0.1250 | +0.0336 |
|  | 3 | base | 0.8672 | 0.7727 | 0.6170 | 0.9074 | 0.7886 | 0.9235 | 0.5156 | 0.7483 |
|  |  | Centiloid | -0.2109 | -0.0455 | +0.1915 | +0.0185 | -0.0302 | -0.1412 | -0.0078 | -0.0839 |
|  | 4 | base | 0.8359 | 0.7273 | 0.5426 | 0.8889 | 0.7450 | 0.9176 | 0.5938 | 0.7785 |
|  |  | Centiloid | -0.2344 | 0.0000 | +0.2021 | +0.0370 | -0.0302 | -0.1529 | +0.1641 | -0.0168 |
|  | 5 | base | 0.7969 | 0.7273 | 0.4894 | 0.8704 | 0.7081 | 0.9353 | 0.4922 | 0.7450 |
|  |  | Centiloid | -0.2812 | 0.0000 | +0.0106 | -0.0185 | -0.1208 | -0.2471 | +0.2344 | -0.0403 |
| PPV | 1 | base | 0.1364 | 0.3333 | 0.0882 | 0.6000 | 0.1562 | 0.5000 | 0.1489 | 0.1887 |
|  |  | Centiloid | -0.0823 | +0.1667 | 0.0000 | -0.1000 | -0.0423 | -0.3000 | -0.0031 | -0.0243 |
|  | 2 | base | 0.5676 | 0.5625 | 0.2593 | 0.6667 | 0.4370 | 0.6875 | 0.2533 | 0.3297 |
|  |  | Centiloid | -0.1457 | +0.0257 | +0.1407 | +0.0606 | +0.0331 | -0.0949 | +0.0570 | +0.0703 |
|  | 3 | base | 0.7018 | 0.7727 | 0.4098 | 0.7222 | 0.6013 | 0.7547 | 0.3800 | 0.5098 |
|  |  | Centiloid | -0.1800 | -0.0455 | +0.0902 | +0.0111 | -0.0376 | -0.1175 | -0.0100 | -0.0049 |
|  | 4 | base | 0.7273 | 0.7692 | 0.4342 | 0.6667 | 0.6142 | 0.7910 | 0.4409 | 0.5875 |
|  |  | Centiloid | -0.1320 | -0.0092 | +0.0858 | +0.0476 | -0.0096 | -0.1136 | +0.0748 | +0.0348 |
|  | 5 | base | 0.7234 | 0.8065 | 0.4146 | 0.6667 | 0.6184 | 0.8358 | 0.3868 | 0.5607 |
|  |  | Centiloid | -0.1367 | -0.0287 | -0.0172 | -0.0145 | -0.0609 | -0.1590 | +0.1271 | +0.0664 |
| NPV | 1 | base | 0.9820 | 0.8000 | 0.9844 | 0.9286 | 0.9531 | 0.9382 | 0.9888 | 0.9551 |
|  |  | Centiloid | -0.0132 | +0.0333 | 0.0000 | -0.0163 | -0.0071 | +0.0052 | -0.0001 | +0.0045 |
|  | 2 | base | 0.8421 | 0.8333 | 0.9474 | 0.9804 | 0.8919 | 0.7783 | 0.9600 | 0.8258 |
|  |  | Centiloid | +0.0164 | +0.0490 | -0.0288 | +0.0004 | +0.0123 | +0.0127 | -0.0035 | +0.0172 |
|  | 3 | base | 0.7603 | 0.8500 | 0.8657 | 0.9245 | 0.8217 | 0.7136 | 0.9296 | 0.7663 |
|  |  | Centiloid | -0.0035 | -0.0500 | -0.0396 | -0.0317 | -0.0116 | +0.0641 | -0.0141 | +0.0519 |
|  | 4 | base | 0.7329 | 0.8000 | 0.8500 | 0.8727 | 0.7900 | 0.6783 | 0.8636 | 0.7296 |
|  |  | Centiloid | +0.0609 | -0.0381 | -0.0360 | -0.0253 | +0.0199 | +0.0732 | -0.0346 | +0.0532 |
|  | 5 | base | 0.7034 | 0.8000 | 0.8364 | 0.8246 | 0.7617 | 0.6285 | 0.7975 | 0.6687 |
|  |  | Centiloid | +0.0381 | -0.1333 | -0.0398 | +0.0118 | +0.0092 | +0.1215 | +0.0255 | +0.1120 |

**Supplementary Table 3.** ROC-AUC of the full and nested models, tested on each leave-out site and time-to-progression. For the full model (amyloid + volume + non-imaging), the ROC-AUC is reported. For each nested model, the difference in ROC-AUC relative to the full model is reported.

|  |  | **leave-out site** | | | | | | |
| --- | --- | --- | --- | --- | --- | --- | --- | --- |
| **progression time-horizon (yrs)** | **feature subset** | **A4** | **ADNI** | **AIBL** | **BLSA** | **HABS** | **MCSA** | **OASIS** |
| 1 | amyloid + volume + non-imaging | 0.8063 | 0.8030 | - | - | 0.2500 | 0.8298 | 0.9074 |
|  | amyloid + non-imaging | -0.1266 | +0.0530 | - | - | +0.4167 | -0.2128 | -0.0344 |
|  | volume + non-imaging | +0.0703 | -0.0076 | - | - | 0.0000 | +0.0160 | +0.0079 |
|  | amyloid + volume | +0.0031 | +0.0152 | - | - | 0.0000 | +0.0186 | +0.0132 |
|  | amyloid | -0.1953 | +0.0227 | - | - | +0.3333 | -0.1995 | -0.0185 |
|  | volume | +0.0734 | -0.0076 | - | - | 0.0000 | +0.0266 | +0.0079 |
|  | non-imaging | -0.0172 | -0.0602 | - | - | 0.0000 | -0.3670***** | -0.0741 |
| 2 | amyloid + volume + non-imaging | 0.7487 | 0.7576 | 1.0000 | 0.7500 | 0.2143 | 0.8004 | 0.9630 |
|  | amyloid + non-imaging | -0.1205 | +0.0492 | -0.0804 | -0.0577 | +0.1429 | -0.0701 | -0.0823 |
|  | volume + non-imaging | +0.0327 | -0.0076 | -0.0714 | 0.0000 | +0.1429 | +0.0394 | -0.0062 |
|  | amyloid + volume | +0.0004 | +0.0227 | -0.0357 | 0.0000 | 0.0000 | -0.0638 | -0.0021 |
|  | amyloid | -0.1144 | +0.0682 | -0.0357 | -0.1538 | +0.1429 | -0.1020 | -0.0823 |
|  | volume | +0.0326 | -0.0227 | -0.0625 | 0.0000 | +0.1429 | +0.0357 | -0.0082 |
|  | non-imaging | -0.1025 | -0.1212 | -0.3929 | +0.1538 | +0.1786 | -0.0494 | -0.0844 |
| 3 | amyloid + volume + non-imaging | 0.7230 | 0.8364 | 0.9881 | 0.6615 | 0.3750 | 0.7078 | 0.8758 |
|  | amyloid + non-imaging | -0.0549 | +0.0227 | -0.0179 | +0.0615 | +0.0625 | +0.0416 | -0.0479 |
|  | volume + non-imaging | +0.0319 | -0.0705 | -0.0655 | 0.0000 | -0.0312 | -0.0119 | -0.0730 |
|  | amyloid + volume | +0.0065 | -0.0023 | +0.0060 | -0.0154 | 0.0000 | -0.0013 | -0.0033 |
|  | amyloid | -0.0643 | -0.0045 | -0.0119 | -0.0154 | +0.1250 | +0.0238 | -0.0403 |
|  | volume | +0.0284 | -0.0773 | -0.0476 | -0.0308 | -0.1562 | -0.0401 | -0.0556 |
|  | non-imaging | -0.0508 | -0.1318 | -0.3750 | +0.2308 | +0.0625 | -0.0091 | -0.1013 |
| 4 | amyloid + volume + non-imaging | 0.7573 | 0.8049 | 0.9603 | 0.7473 | 0.4583 | 0.7538 | 0.8489 |
|  | amyloid + non-imaging | -0.0299 | +0.0625 | +0.0317 | -0.0440 | +0.0625 | -0.0590 | -0.0419 |
|  | volume + non-imaging | -0.0015 | -0.0966 | -0.1468 | -0.0659 | -0.1458 | -0.0562 | -0.2232 |
|  | amyloid + volume | +0.0009 | -0.0019 | -0.0040 | -0.0110 | 0.0000 | +0.0005 | 0.0000 |
|  | amyloid | -0.0551 | +0.0455 | +0.0238 | -0.1209 | +0.1250 | -0.0431 | -0.0409 |
|  | volume | -0.0035 | -0.1004 | -0.1429 | -0.1319 | -0.1250 | -0.0317 | -0.1033 |
|  | non-imaging | -0.0837 | -0.1042 | -0.3532 | +0.1868 | +0.0208 | -0.0486 | -0.1345 |
| 5 | amyloid + volume + non-imaging | 0.7478 | 0.7743 | 0.8622 | 0.7902 | 0.5000 | 0.7640 | 0.8395 |
|  | amyloid + non-imaging | -0.0323 | +0.0721 | +0.0102 | -0.0210 | 0.0000 | -0.0401 | -0.0463 |
|  | volume + non-imaging | +0.0039 | -0.1144 | -0.1097 | -0.1678 | -0.1667 | -0.0381 | -0.1906 |
|  | amyloid + volume | +0.0008 | -0.0047 | -0.0179 | 0.0000 | +0.0167 | -0.0829 | -0.0031 |
|  | amyloid | -0.0480 | +0.0533 | -0.0077 | -0.0490 | +0.0333 | -0.0490 | -0.0633 |
|  | volume | +0.0010 | -0.1097 | -0.1148 | -0.1608 | -0.1667 | -0.0416 | +0.0008 |
|  | non-imaging | -0.0602 | -0.1003 | -0.2423 | +0.0105 | -0.0333 | -0.0482 | -0.1196 |
| **p < 0.05* | | | | | | | | |

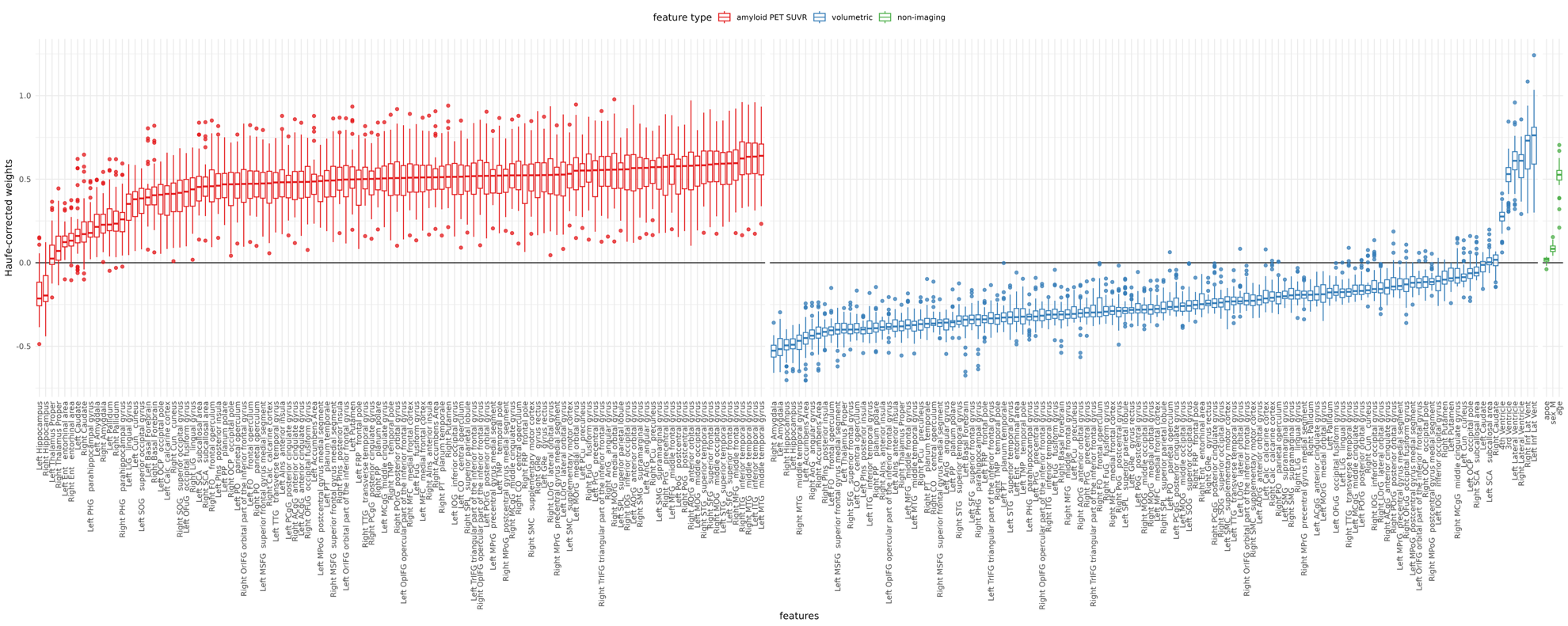
**Supplementary Figure 2.** Distributions of Haufe covariance-corrected linear SVM weights of each input feature, aggregated across all leave-out groups (site & tracer) and all times-to-progression. Red plots indicate amyloid PET SUVR features, blue plots indicate volumetric features, and green plots indicate non-imaging features.

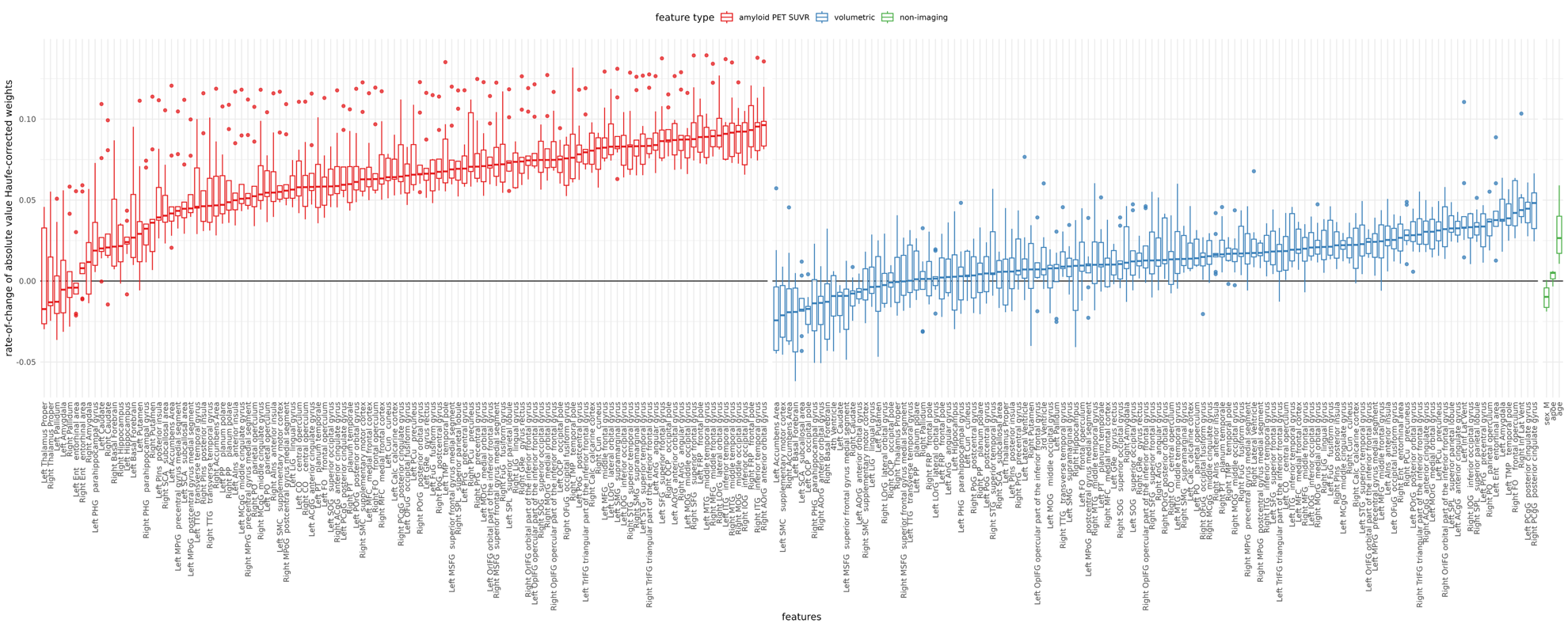
**Supplementary Figure 3.** Distributions of the slope of the magnitude of covariance-corrected weights versus times-to-progression, aggregated across all leave-out groups (site & tracer) and all times-to-progression. Red plots indicate amyloid PET SUVR features, blue plots indicate volumetric features, and green plots indicate non-imaging features.

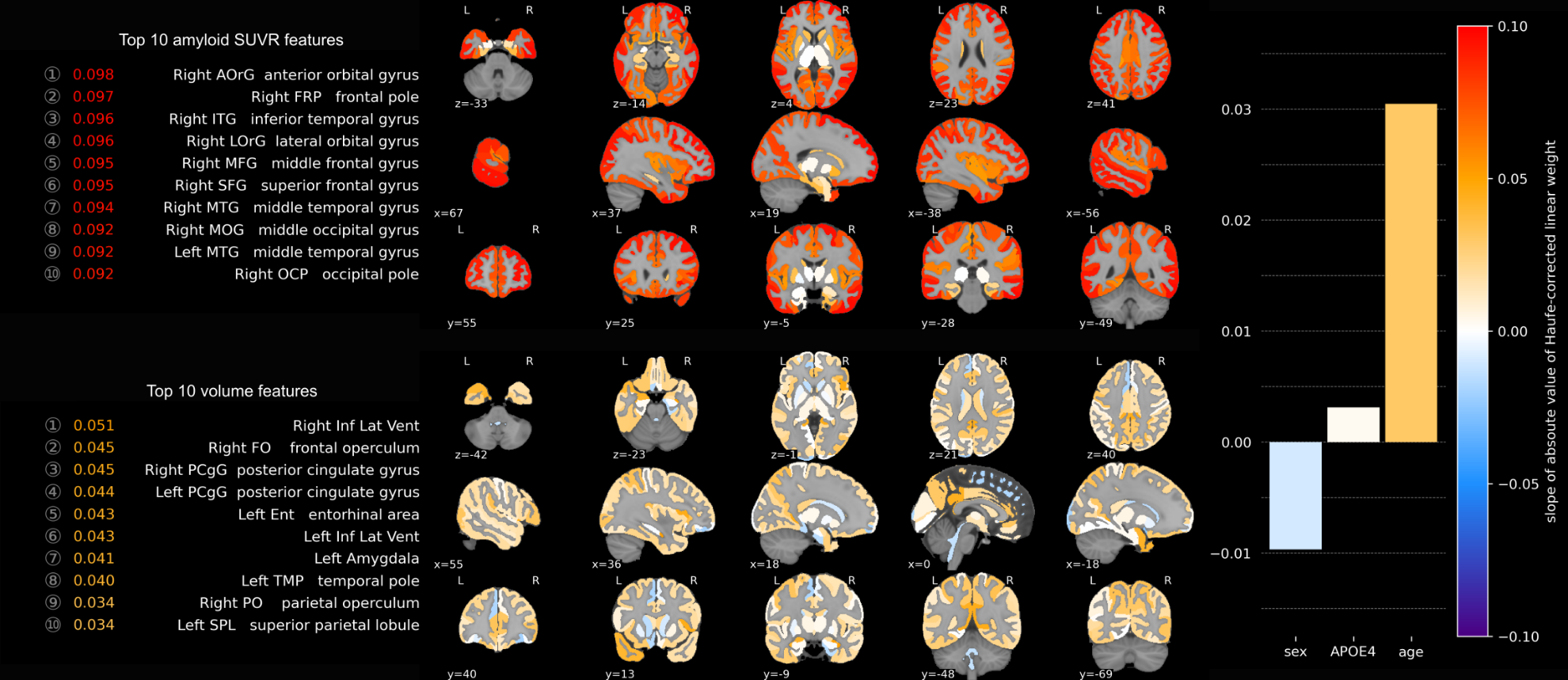

**Supplementary Figure 4.** Mean slope of the magnitude of covariance-corrected SVM weights versus times-to-progression, aggregated across all 45 trained models.

**Supplementary Table 4.** Natural cubic spline modeling of longitudinal PACC of predicted stables, predicted progressors, and the overall A4 cohort. Model-adjusted PACC distributions estimated at 240-weeks is displayed for each predicted class. The contrast in model-adjusted PACC between predicted class and its statistical significance is also shown.

| **progression time-horizon (yrs)** | **predicted class** | **n** | **model-adjusted mean PACC (± SE)** | **lower 95% CI** | **upper 95% CI** | **effect size (± SE)** | **p-value (Bonferroni-adjusted)** |
| --- | --- | --- | --- | --- | --- | --- | --- |
| **Solanezumab** | | | | | | | |
| 1 | overall | 404 | -2.79 ± 0.484 | -3.735 | -1.839 |  |  |
|  | progressor | 61 | -4.04 ± 0.896 | -5.800 | -2.285 |  |  |
|  | stable | 343 | -1.53 ± 0.283 | -2.086 | -0.9763 |  |  |
|  | stable - progressor | 404 | 2.51 ± 0.912 | 0.7235 | 4.300 | 0.83 ± 0.302 | 0.06 |
| 2 | overall | 404 | -2.61 ± 0.373 | -3.343 | -1.882 |  |  |
|  | progressor | 110 | -4.12 ± 0.656 | -5.404 | -2.830 |  |  |
|  | stable | 294 | -1.11 ± 0.274 | -1.646 | -0.5710 |  |  |
|  | stable - progressor | 404 | 3.01 ± 0.676 | 1.684 | 4.333 | 1 ± 0.225 | <0.001*** |
| 3 | overall | 404 | -2.39 ± 0.325 | -3.026 | -1.750 |  |  |
|  | progressor | 140 | -3.89 ± 0.546 | -4.961 | -2.822 |  |  |
|  | stable | 264 | -0.884 ± 0.277 | -1.428 | -0.3408 |  |  |
|  | stable - progressor | 404 | 3.01 ± 0.571 | 1.888 | 4.126 | 1 ± 0.191 | <0.001*** |
| 4 | overall | 404 | -2.3 ± 0.32 | -2.932 | -1.676 |  |  |
|  | progressor | 150 | -3.74 ± 0.534 | -4.789 | -2.694 |  |  |
|  | stable | 254 | -0.867 ± 0.277 | -1.409 | -0.3242 |  |  |
|  | stable - progressor | 404 | 2.88 ± 0.56 | 1.778 | 3.972 | 0.958 ± 0.187 | <0.001*** |
| 5 | overall | 404 | -2.38 ± 0.332 | -3.031 | -1.728 |  |  |
|  | progressor | 143 | -3.92 ± 0.568 | -5.032 | -2.804 |  |  |
|  | stable | 261 | -0.84 ± 0.264 | -1.358 | -0.3231 |  |  |
|  | stable - progressor | 404 | 3.08 ± 0.586 | 1.929 | 4.227 | 1.03 ± 0.196 | <0.001*** |
| **Placebo** | | | | | | | |
| 1 | overall | 444 | -2.51 ± 0.347 | -3.188 | -1.826 |  |  |
|  | progressor | 72 | -3.6 ± 0.624 | -4.826 | -2.373 |  |  |
|  | stable | 372 | -1.41 ± 0.23 | -1.866 | -0.9629 |  |  |
|  | stable - progressor | 444 | 2.19 ± 0.635 | 0.9398 | 3.431 | 0.812 ± 0.236 | 0.006** |
| 2 | overall | 444 | -2.37 ± 0.277 | -2.914 | -1.827 |  |  |
|  | progressor | 132 | -3.77 ± 0.47 | -4.688 | -2.843 |  |  |
|  | stable | 312 | -0.975 ± 0.219 | -1.406 | -0.5452 |  |  |
|  | stable - progressor | 444 | 2.79 ± 0.481 | 1.847 | 3.733 | 1.04 ± 0.18 | <0.001*** |
| 3 | overall | 444 | -2.06 ± 0.258 | -2.561 | -1.550 |  |  |
|  | progressor | 158 | -3.12 ± 0.419 | -3.942 | -2.297 |  |  |
|  | stable | 286 | -0.992 ± 0.223 | -1.429 | -0.5548 |  |  |
|  | stable - progressor | 444 | 2.13 ± 0.431 | 1.283 | 2.972 | 0.794 ± 0.161 | <0.001*** |
| 4 | overall | 444 | -2.2 ± 0.261 | -2.714 | -1.690 |  |  |
|  | progressor | 149 | -3.51 ± 0.429 | -4.349 | -2.667 |  |  |
|  | stable | 295 | -0.896 ± 0.223 | -1.333 | -0.4589 |  |  |
|  | stable - progressor | 444 | 2.61 ± 0.441 | 1.748 | 3.476 | 0.977 ± 0.165 | <0.001*** |
| 5 | overall | 444 | -2.17 ± 0.262 | -2.685 | -1.659 |  |  |
|  | progressor | 150 | -3.39 ± 0.428 | -4.229 | -2.549 |  |  |
|  | stable | 294 | -0.955 ± 0.224 | -1.394 | -0.5160 |  |  |
|  | stable - progressor | 444 | 2.43 ± 0.44 | 1.571 | 3.296 | 0.909 ± 0.165 | <0.001*** |
| **p < 0.05*  ***p < 0.01*  ****p < 0.001* | | | | | | | |

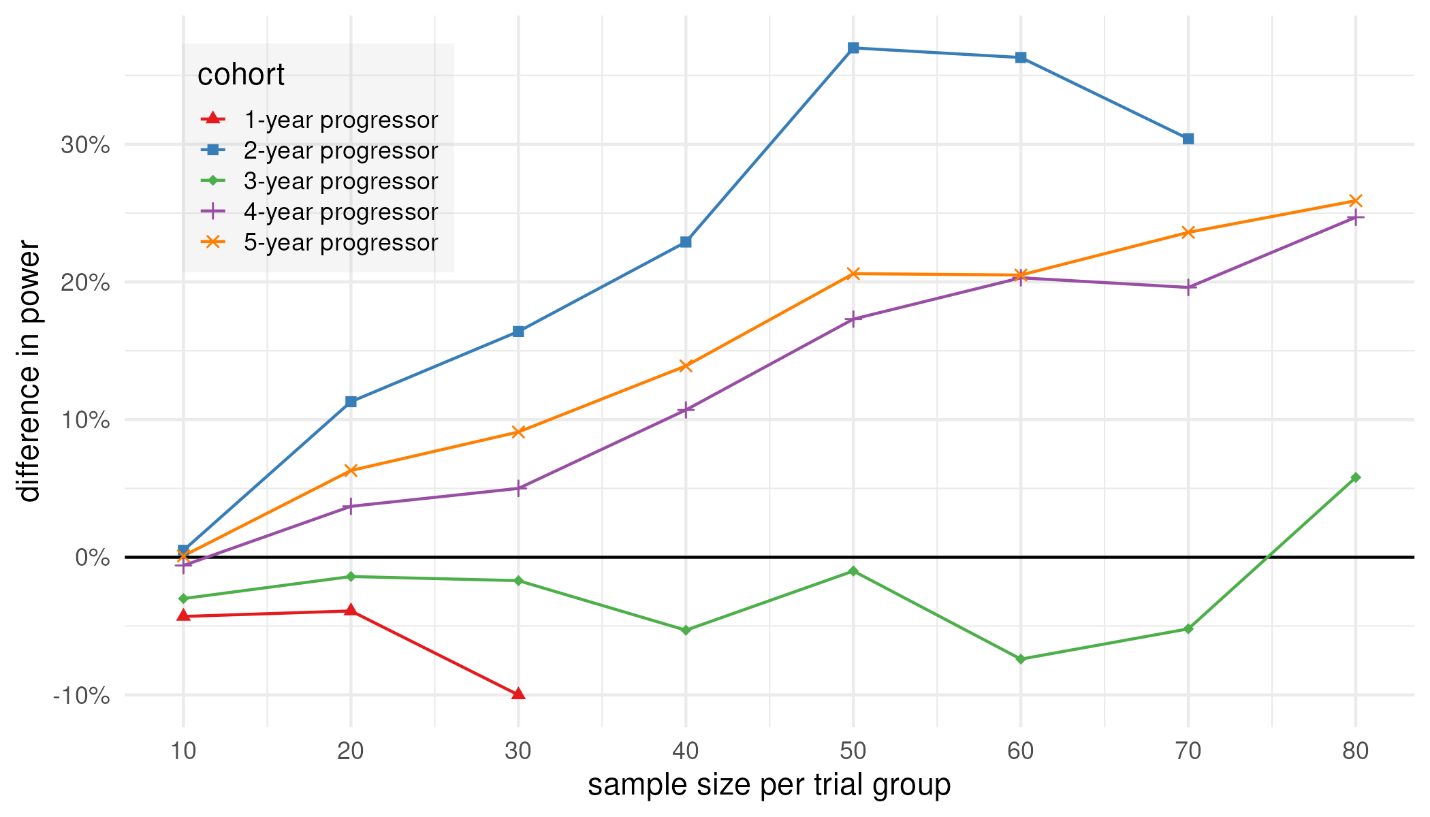

**Supplementary Figure 5.** Difference in power to detect a treatment effect in FBP cortical SUVR between placebo and Solanezumab arms of the A4 study cohort, relative to the original, unenriched cohort.
